# Do school closures and school reopenings affect community transmission of COVID-19? A systematic review of observational studies

**DOI:** 10.1101/2021.01.02.21249146

**Authors:** Sebastian Walsh, Avirup Chowdhury, Vickie Braithwaite, Simon Russell, Jack Birch, Joseph Ward, Claire Waddington, Carol Brayne, Chris Bonell, Russell Viner, Oliver Mytton

## Abstract

**Background:** School closures are associated with significant negative consequences and exacerbate inequalities. They were implemented worldwide to control SARS-CoV-2 in the first half of 2020, but their effectiveness, and the effects of lifting them, remain uncertain. This review summarises observational evidence of the effect of school closures and school reopenings on SARS-CoV-2 community transmission.

**Methods:** The study protocol was registered on Prospero (ID:CRD42020213699). On 07 January 2021 we searched PubMed, Web of Science, Scopus, CINAHL, the WHO Global COVID-19 Research Database, ERIC, the British Education Index, the Australian Education Index, and Google. We included observational studies with quantitative estimates of the effect of school closures/reopenings on SARS-CoV-2 community transmission. We excluded prospective modelling studies and intra-school transmission studies. We performed a narrative synthesis due to data heterogeneity. We used the ROBINS-I tool to assess risk of bias.

**Findings:** We identified 7,474 articles, of which 40 were included, with data from 150 countries. Of these 32 studies assessed school closures, and 11 examined reopenings. There was substantial heterogeneity between school closure studies, with half of the studies at lower risk of bias reporting reduced community transmission by up to 60%, and half reporting null findings. The majority (n=3 out of 4) of school reopening studies at lower risk of bias reported no associated increases in transmission.

**Conclusions:** School closure studies were at risk of confounding and collinearity from other non-pharmacological interventions implemented around the same time as school closures, and the effectiveness of closures remains uncertain. School reopenings, in areas of low transmission and with appropriate mitigation measures, were generally not accompanied by increasing community transmission. With such varied evidence on effectiveness, and the harmful effects, policymakers should take a measured approach before implementing school closures; and should look to reopen schools in times of low transmission, with appropriate mitigation measures.

## Introduction

School closures have been a common strategy to control the spread of SARS-CoV-2 during the COVID-19 pandemic. By 2 April 2020, 172 nations had enacted full closures or partial ‘dismissals’, affecting nearly 1·5 billion children(1). As cases of COVID-19 started to fall, many countries looked to reopen schools, often with significant mitigation measures in place(2). Over the northern hemisphere winter of 2020-21, many countries again closed schools with the aim of controlling a second pandemic wave. School closures have substantial negative consequences for children’s wellbeing and education, which will impact on life chances and long-term health(3, 4). Closures exacerbate existing inequalities, with greater impacts upon more deprived children because those from higher income families have better opportunities for remote learning.

The role of non-pharmaceutical interventions (NPIs) collectively in limiting community spread is established. However, the specific contribution of school closures remains unclear. Observational studies suggest that school-aged children, particularly teenagers, play a role in transmission to peers and bringing infection into households(5), although the relative importance compared to adults remains unclear(6). Younger children appear less susceptible to infection and may play a smaller role in community transmission, compared with older children and adults(7). Although some modelling studies have suggested that school closures can reduce SARS-CoV-2 community transmission(8), others disagree(9, 10).

A rapid systematic review published in April 2020 found a small number of studies of the effectiveness of school closures in controlling the spread of coronaviruses(11). However, this review was undertaken very early in the pandemic and included no observational data on SARS-CoV-2. Since then many studies on the effects of closing or re-opening schools on SARS-CoV-2 community transmission have been published, but there has been no systematic review of these studies. A clearer understanding of the impact of school closures and reopenings on community transmission is essential to aid policymakers in deciding if and when to implement school closures in response to rising virus prevalence, and when it is prudent to reopen schools. Here, we synthesise the observational evidence of the impact of closing or reopening schools on community transmission of SARS-CoV-2.

## Methods

The study protocol for this systematic review is registered on Prospero (ID:CRD42020213699).

### Inclusion and Exclusion Criteria

We included any empirical study which reported a quantitative estimate of the effect of school closure or reopening on community transmission of SARS-CoV-2. We considered ‘school’ to include early years settings (e.g. nurseries or kindergartens), primary schools, and secondary school, but excluded further or higher education (e.g. universities). Community transmission was defined as any measure of community infections rate, hospital admissions, or mortality attributed to COVID-19. We included studies published in 2020 or 2021 only. We included pre-prints, peer-reviewed and grey literature. We did not apply any restriction on language, but all searches were undertaken in English. We excluded prospective modelling studies and studies in which the assessed outcome was exclusively transmission within the school environment rather than the wider community.

### Search strategy

We searched PubMed, Web of Science, Scopus, CINAHL, the WHO Global COVID-19 Research Database (including medRxiv and SSRN), ERIC, the British Education Index, and the Australian Education Index, searching title and abstracts for terms related to SARS-CoV-2 AND terms related to schools or NPIs. To search the grey literature, we searched Google. We also included papers identified through professional networks. Full details of the search strategy are included in Appendix A. Searches were undertaken first on 12 October 2020 and updated on 07 January 2021.

### Data extraction and risk of bias assessment

Article titles and abstracts were imported into the Rayyan QCRI webtool(12). Two reviewers independently screened titles and abstracts, retrieved full texts of potentially relevant articles, and assessed eligibility for inclusion.

Two reviewers independently extracted data and assessed risk of bias. Data extraction was performed using a pre-agreed extraction template which collected information on publication type (peer-reviewed or pre-print), country, study design, exposure type (school closure or reopening), setting type (primary or secondary), study period, unit of observation, confounders adjusted for, other NPIs in place, analysis method, outcome measure, and findings. We used the Cochrane Risk of Bias In Non-randomised Studies of Interventions (ROBINS-I) tool(13) to evaluate bias.

Discrepancies were resolved by discussion in the first instance and by a third reviewer where necessary.

### Data synthesis

Given the heterogeneous nature of the studies, prohibiting meta-analysis, a narrative synthesis was conducted. Schools often reopened with significant COVID-19 infection prevention and control measures in place, meaning that the effect of lifting restrictions may have been different from the effect of imposing them. We therefore considered the studies of school closures and school reopenings separately. We also aimed to evaluate differential effects for primary and secondary schools if data allowed.

## Results

We identified 7,474 studies (Figure 1). After removing 2,339 duplicates, 5,135 unique records were screened for inclusion. We excluded 4,842 records at the title or abstract stage, leaving 293 records for full text review. Of these, 40(14–53) met the inclusion criteria.

**Figure 1:**
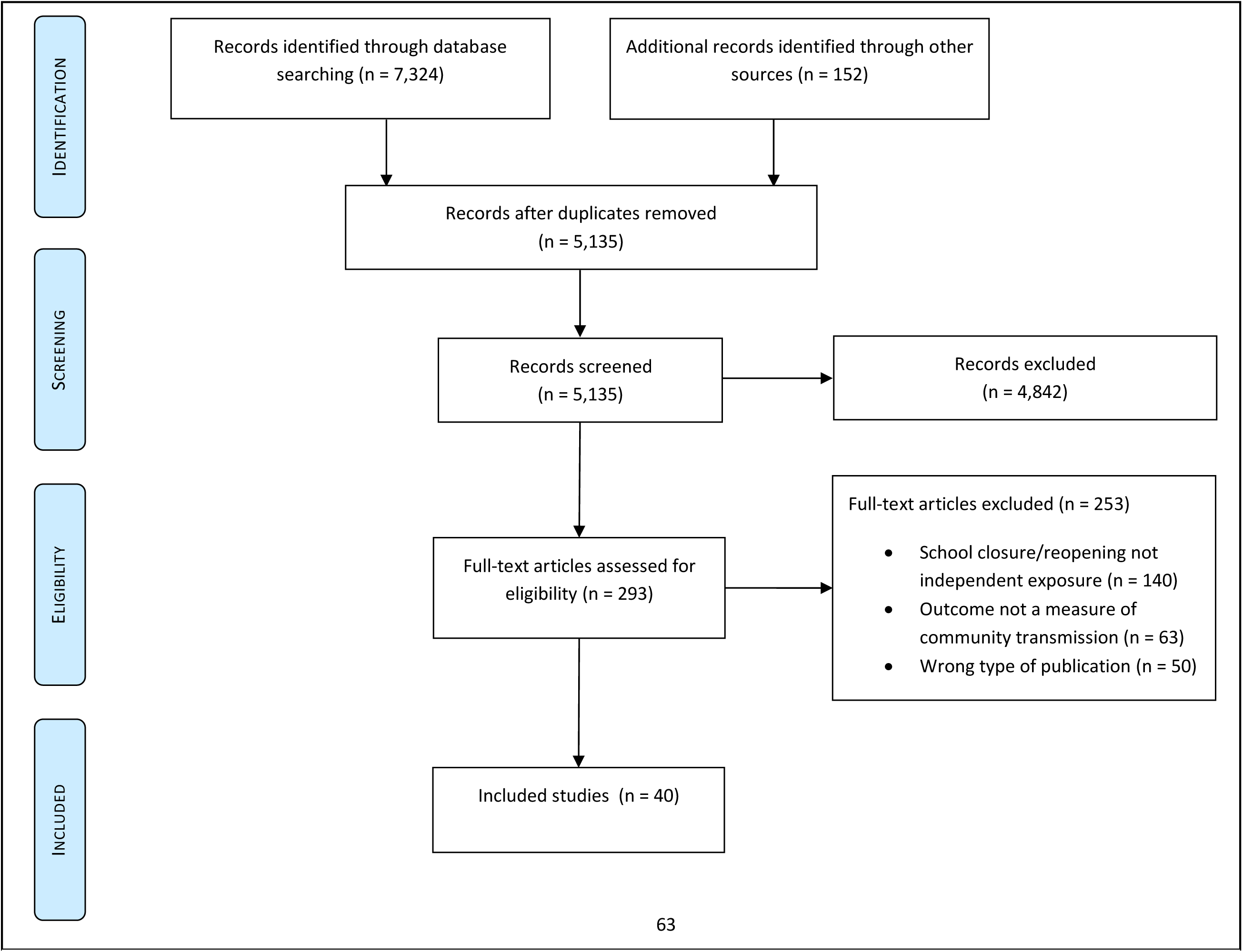
Flow diagram of study selection process.

### Description of studies

Included studies are described in Table 1, grouped by exposure type and study design. Of these, 32 studies(14,15,18–21,23,24,26,29–40,42–44,46–53) reported the effect of school closures on community transmission of SARS-CoV-2, 11(16,22–25,27,28,35,43–45) examined school reopening, and 3(16,17,41) investigated the effect of school holidays. Some studies considered more than one exposure. All studies used data from national Government sources or international data repositories. A total of 15 studies were from peer reviewed journals, whilst 24 studies were from pre-print servers, and one study was a conference abstract.

**Table 1:**
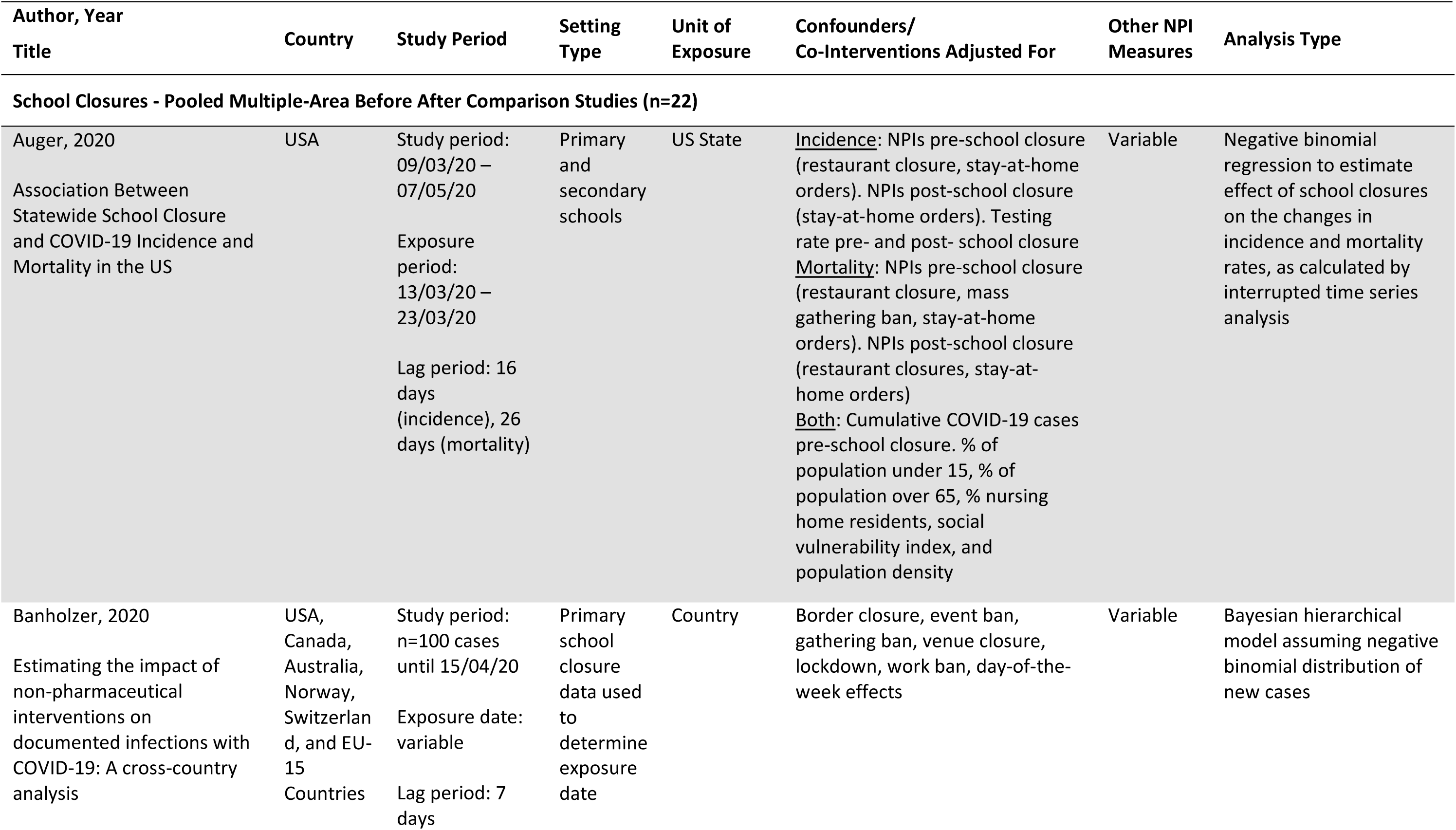
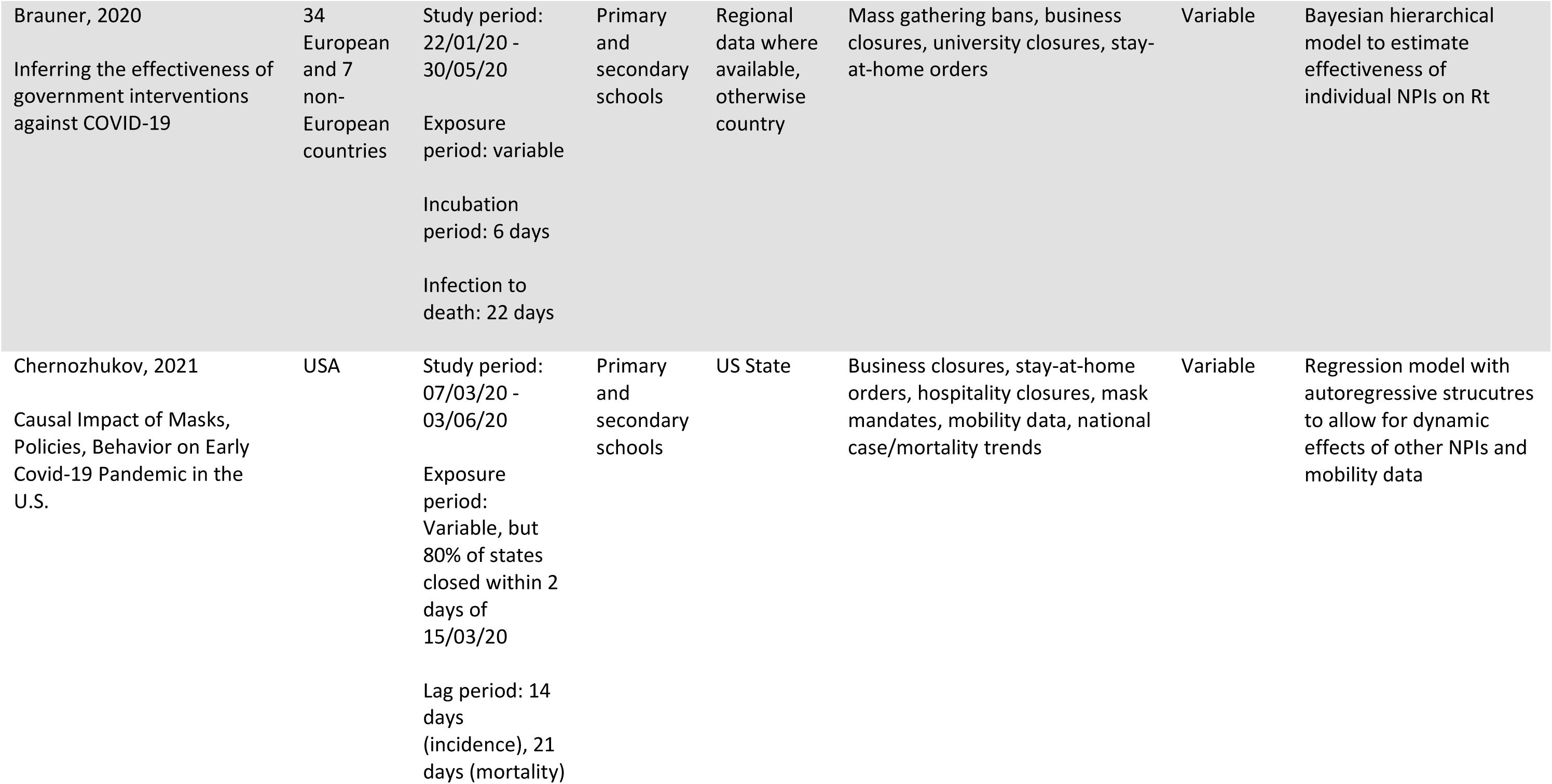
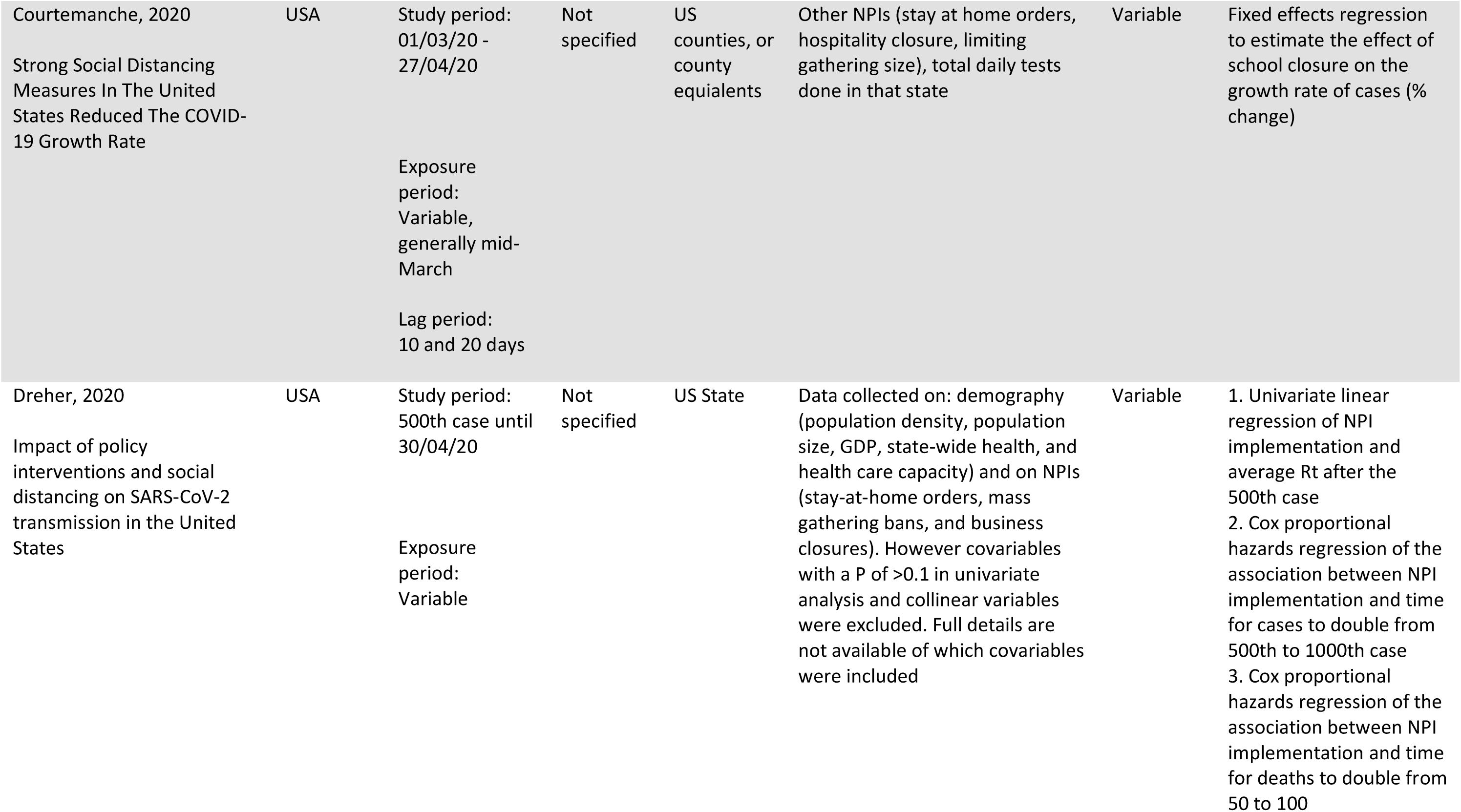
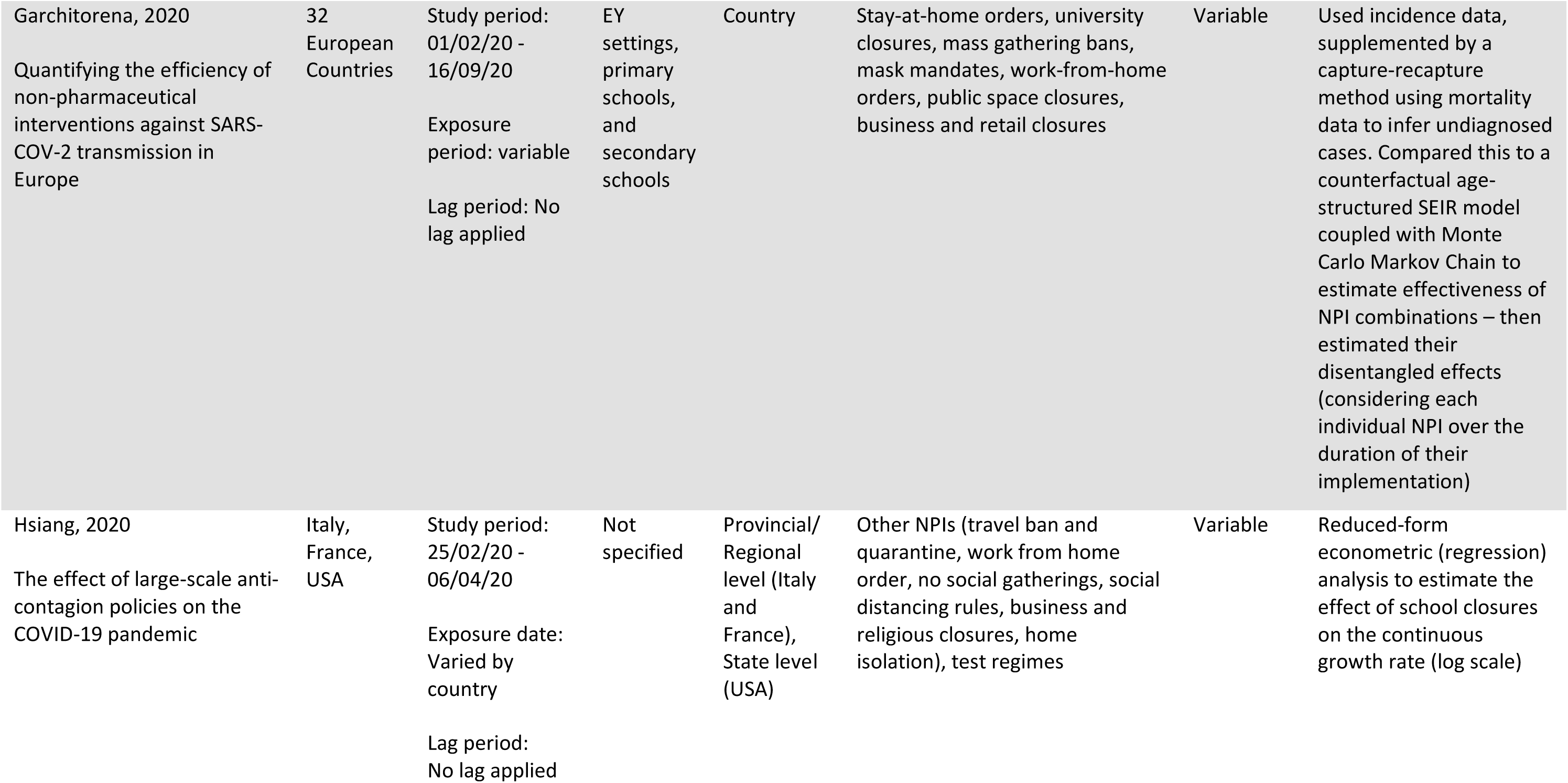
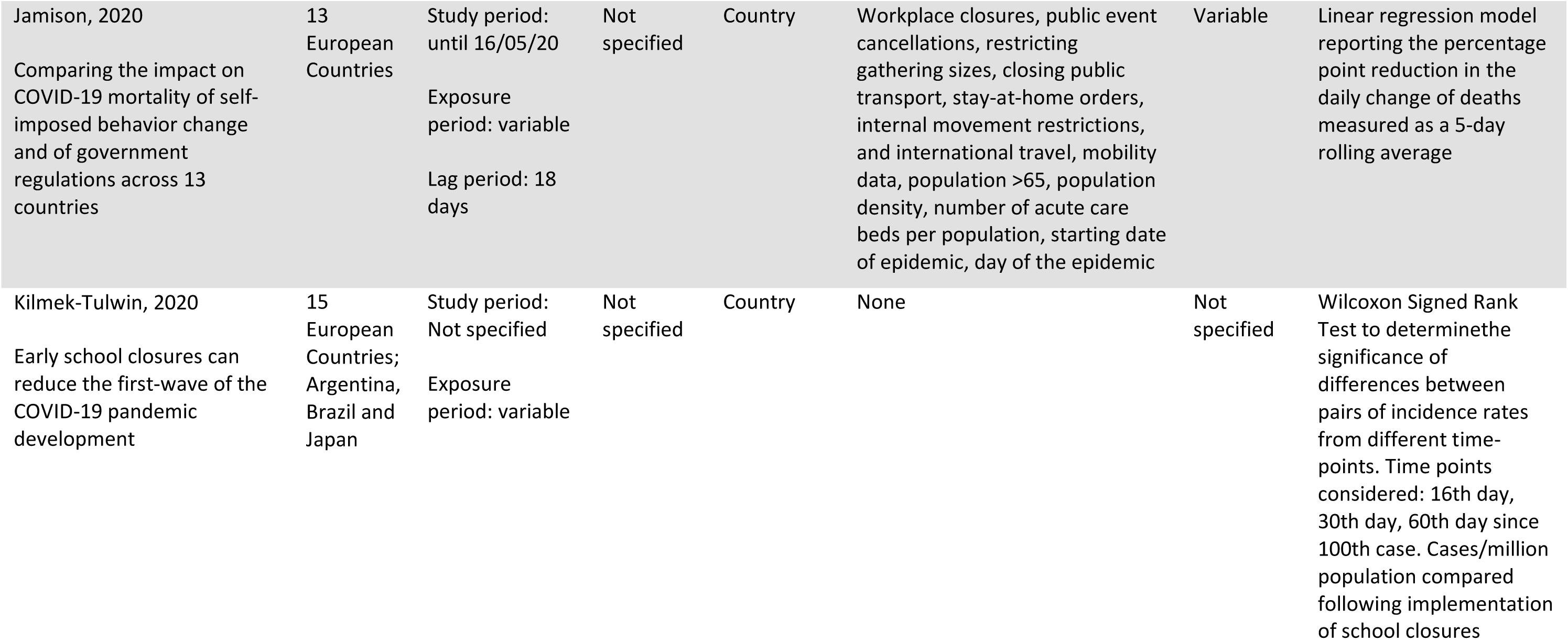
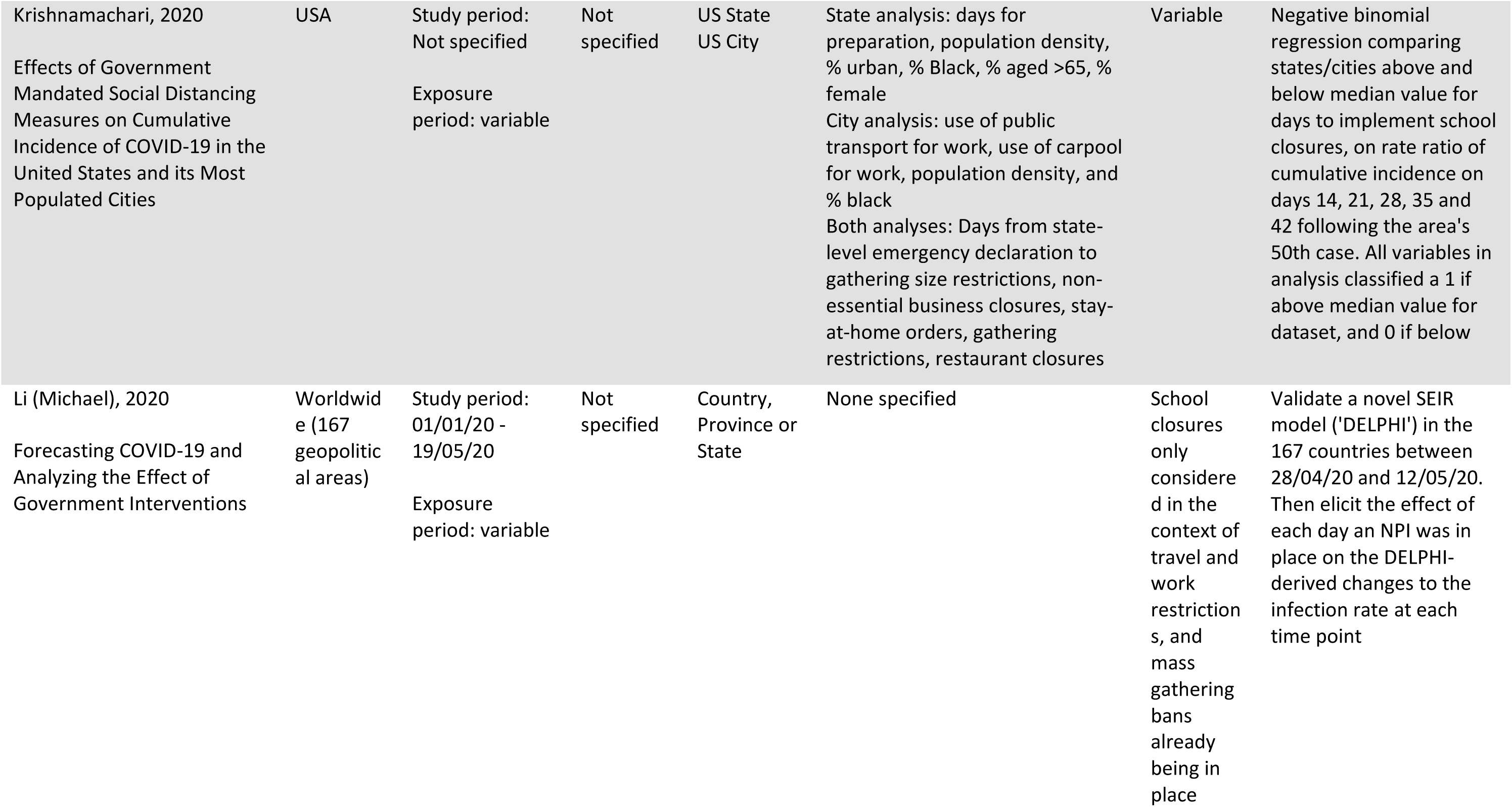
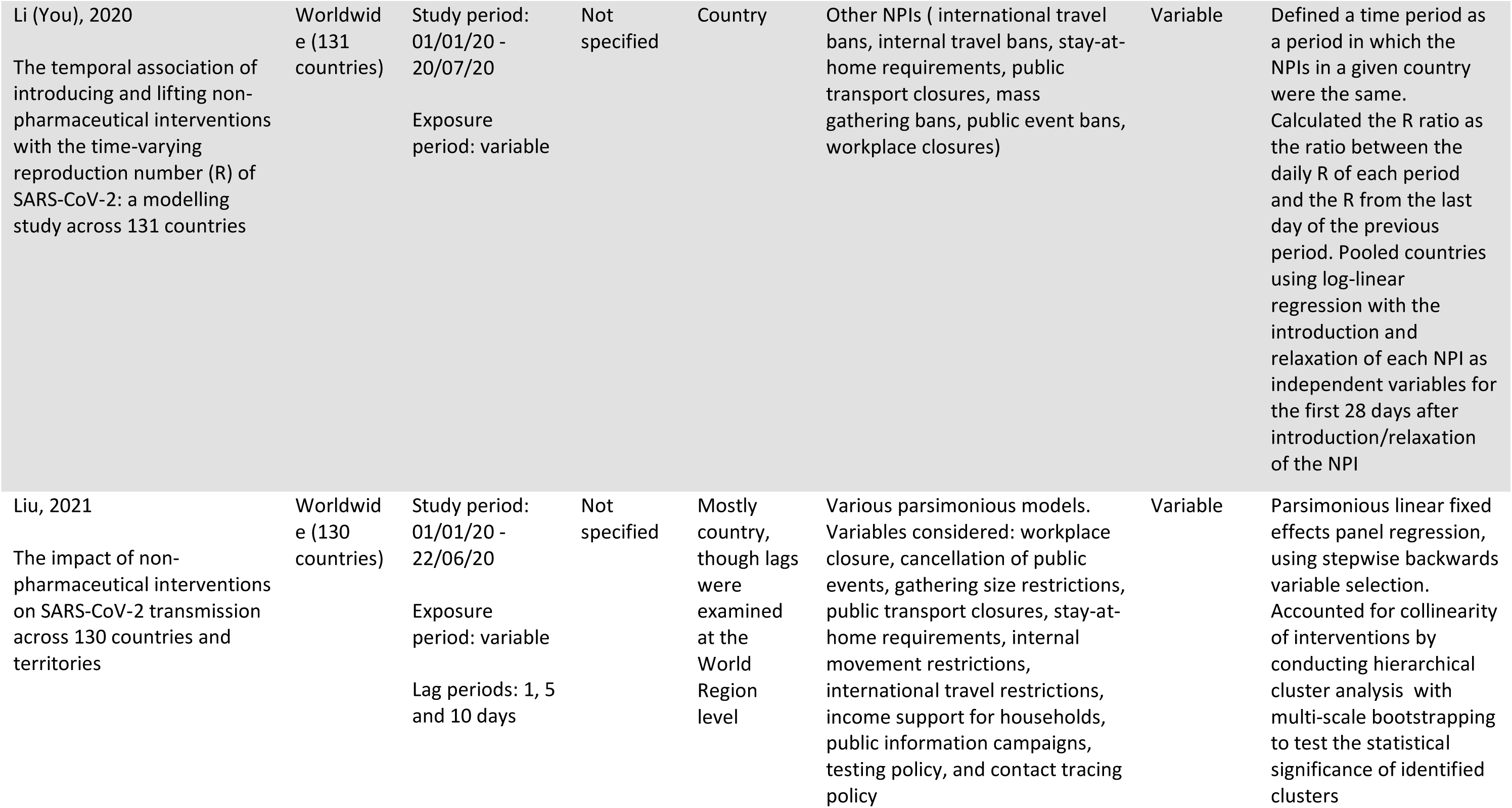
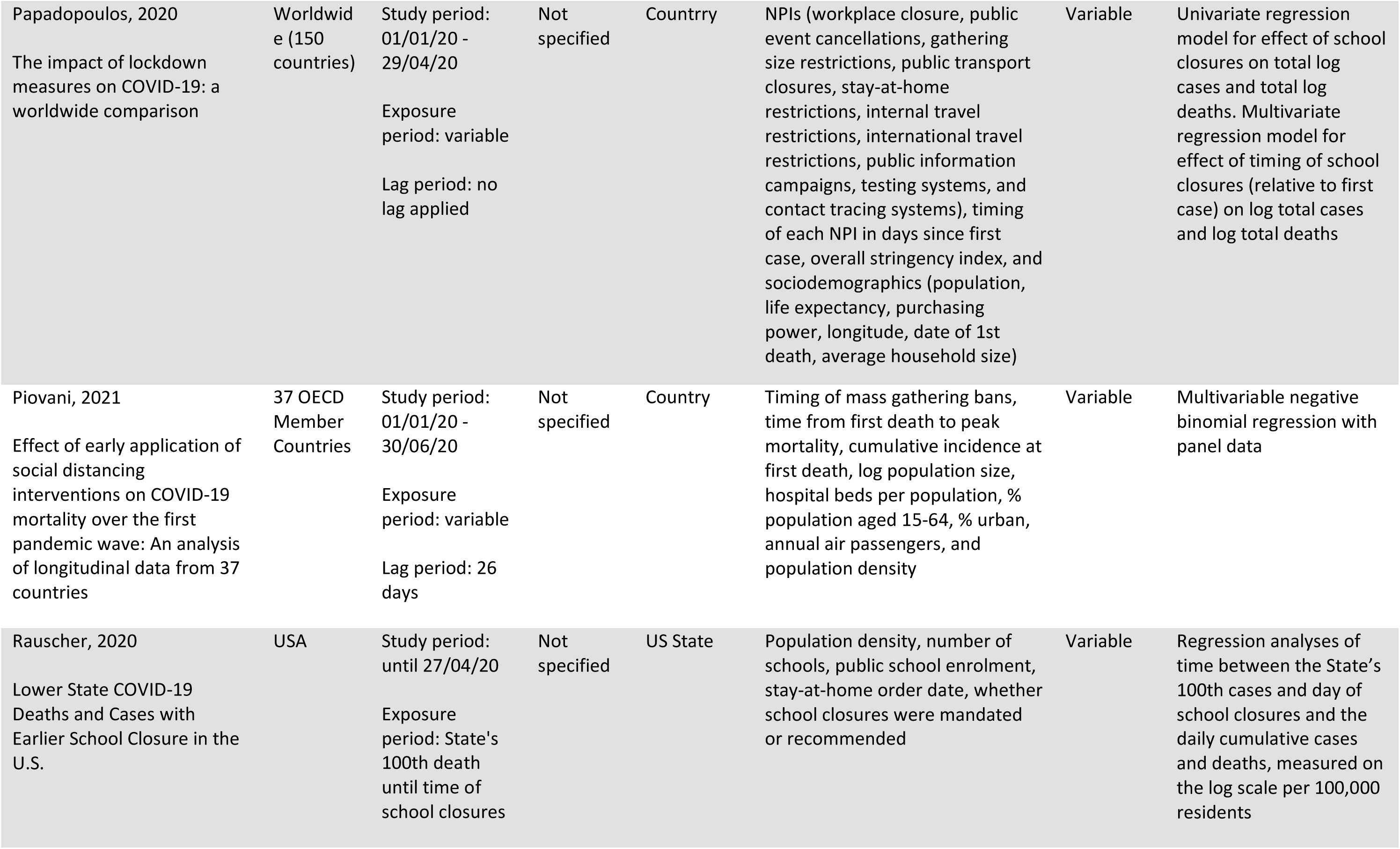
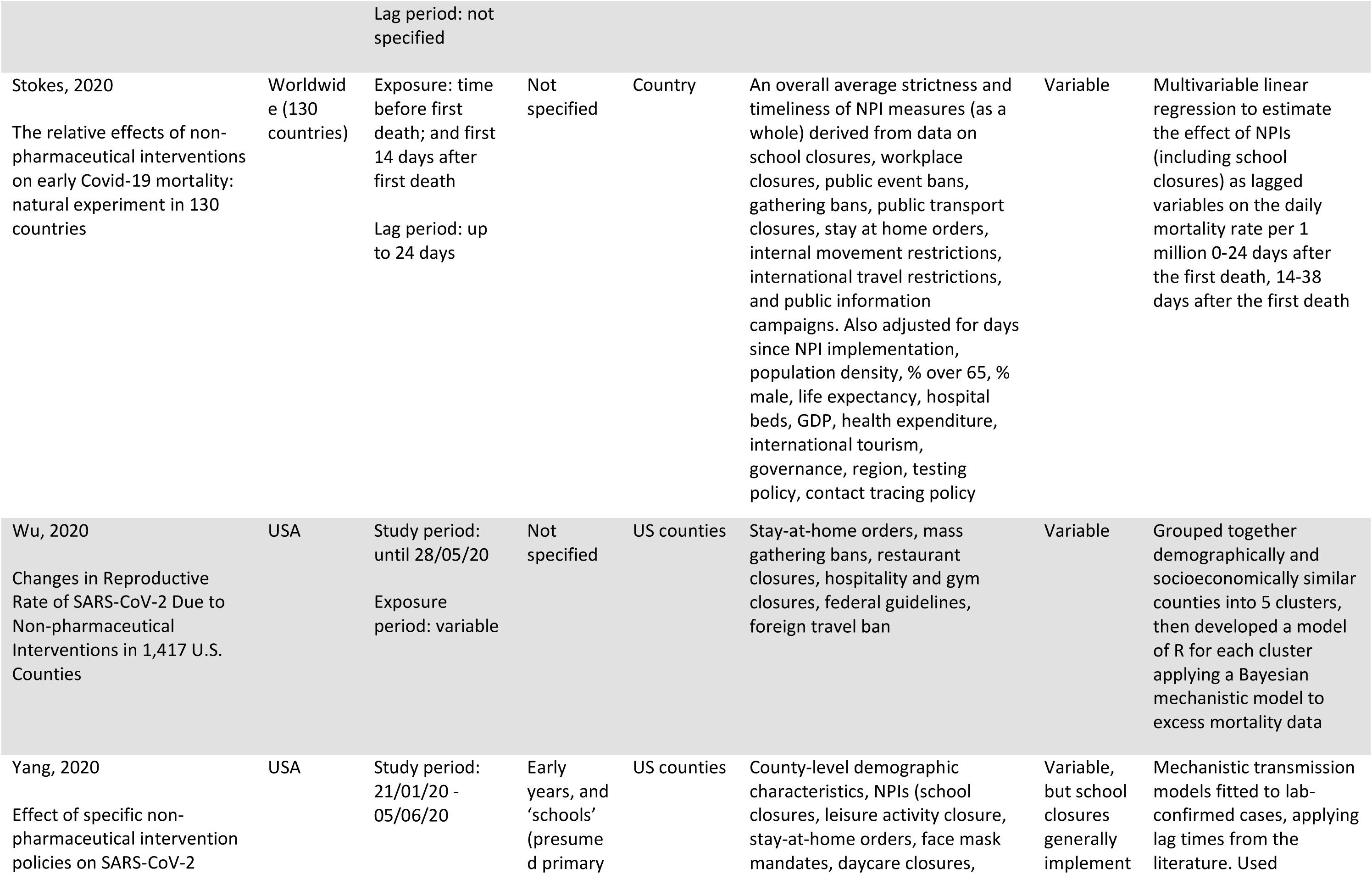
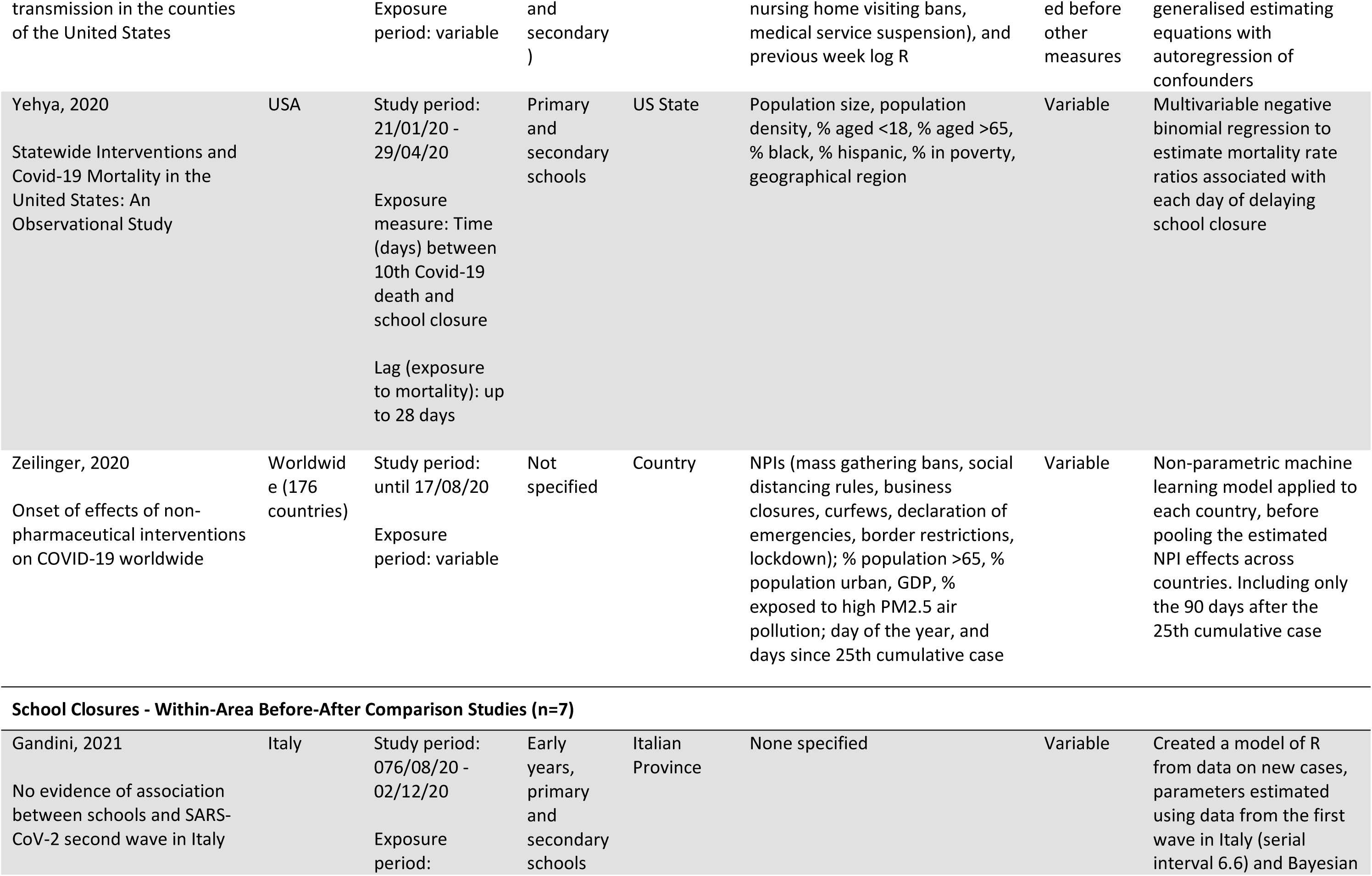
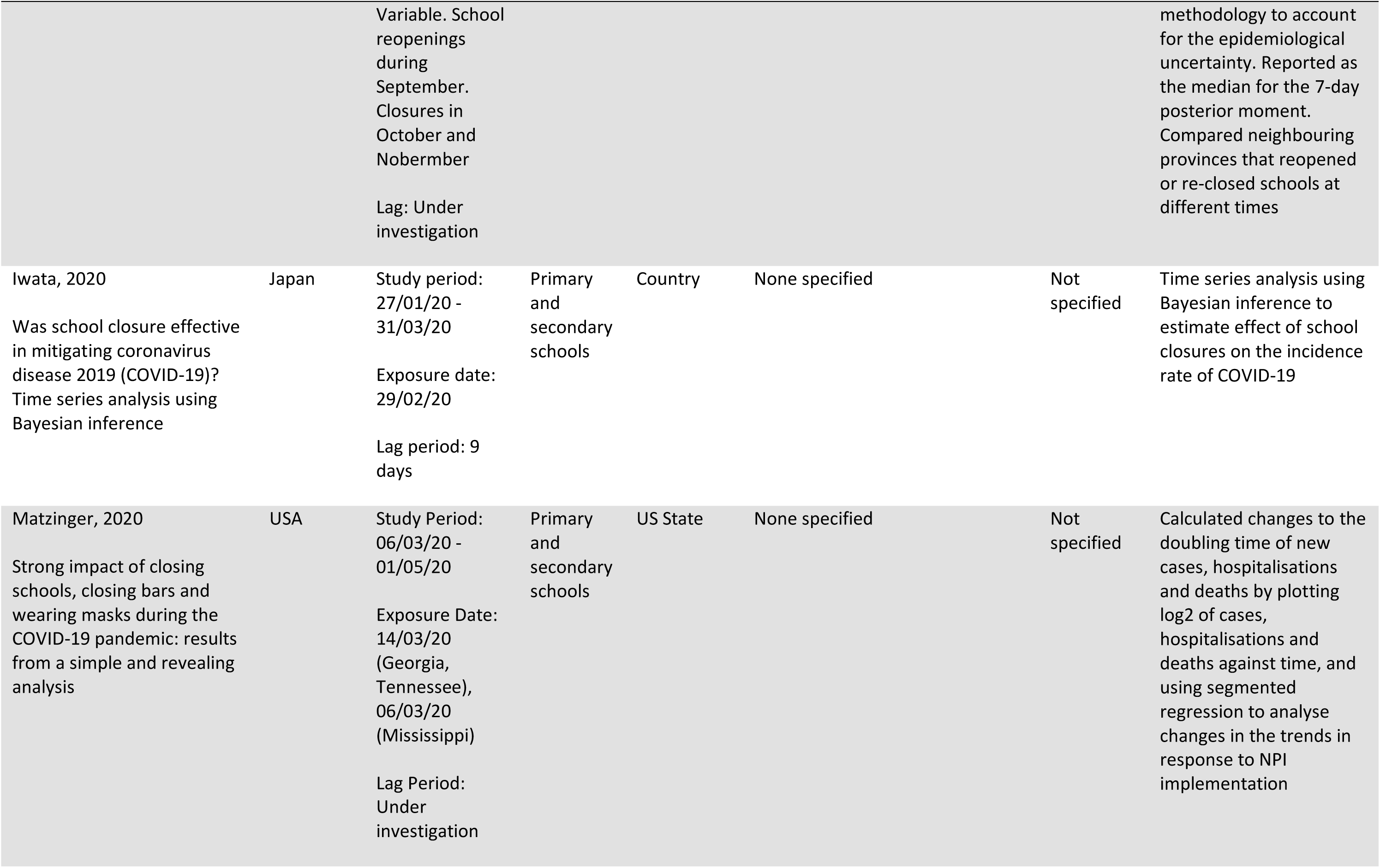
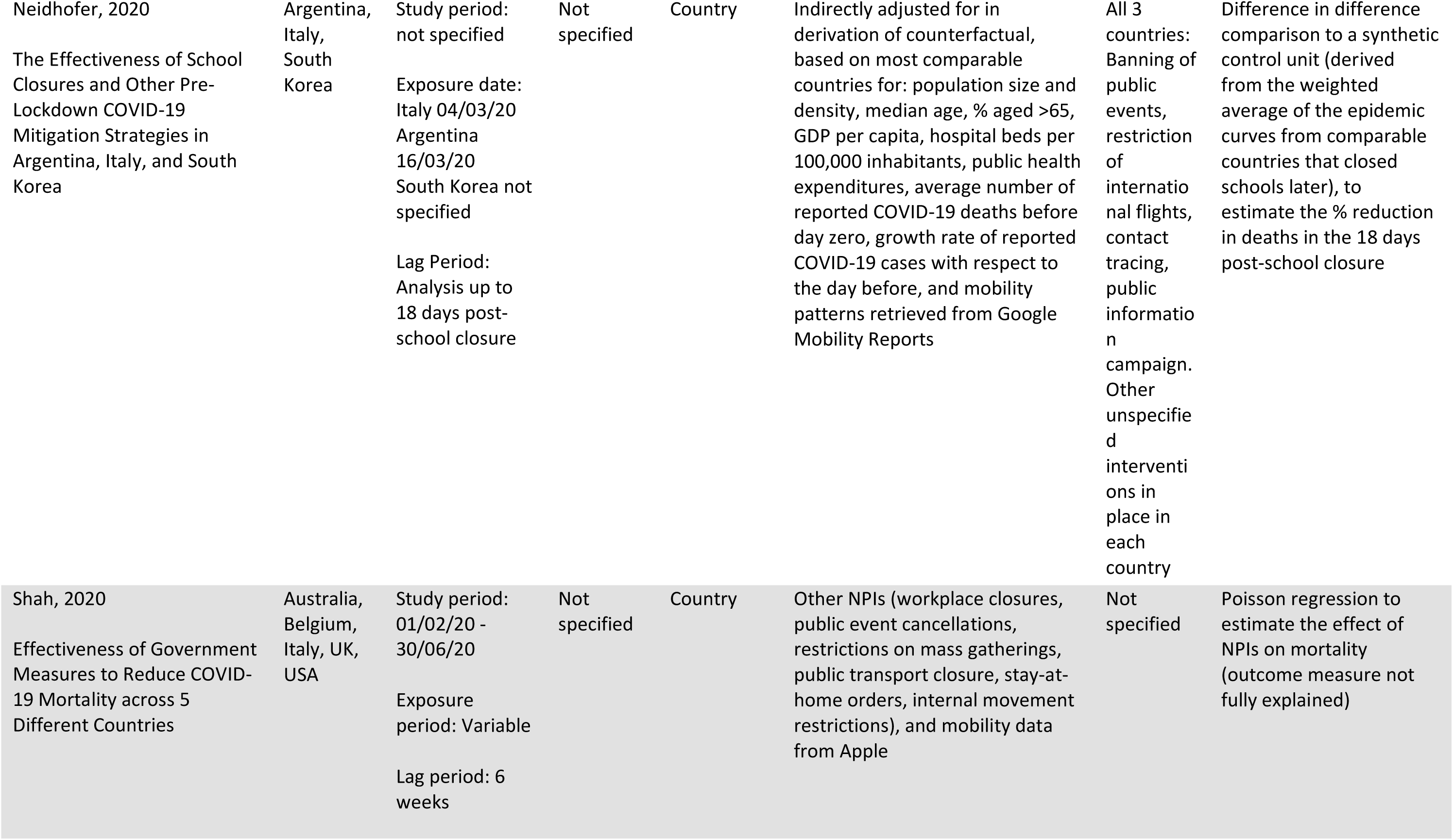
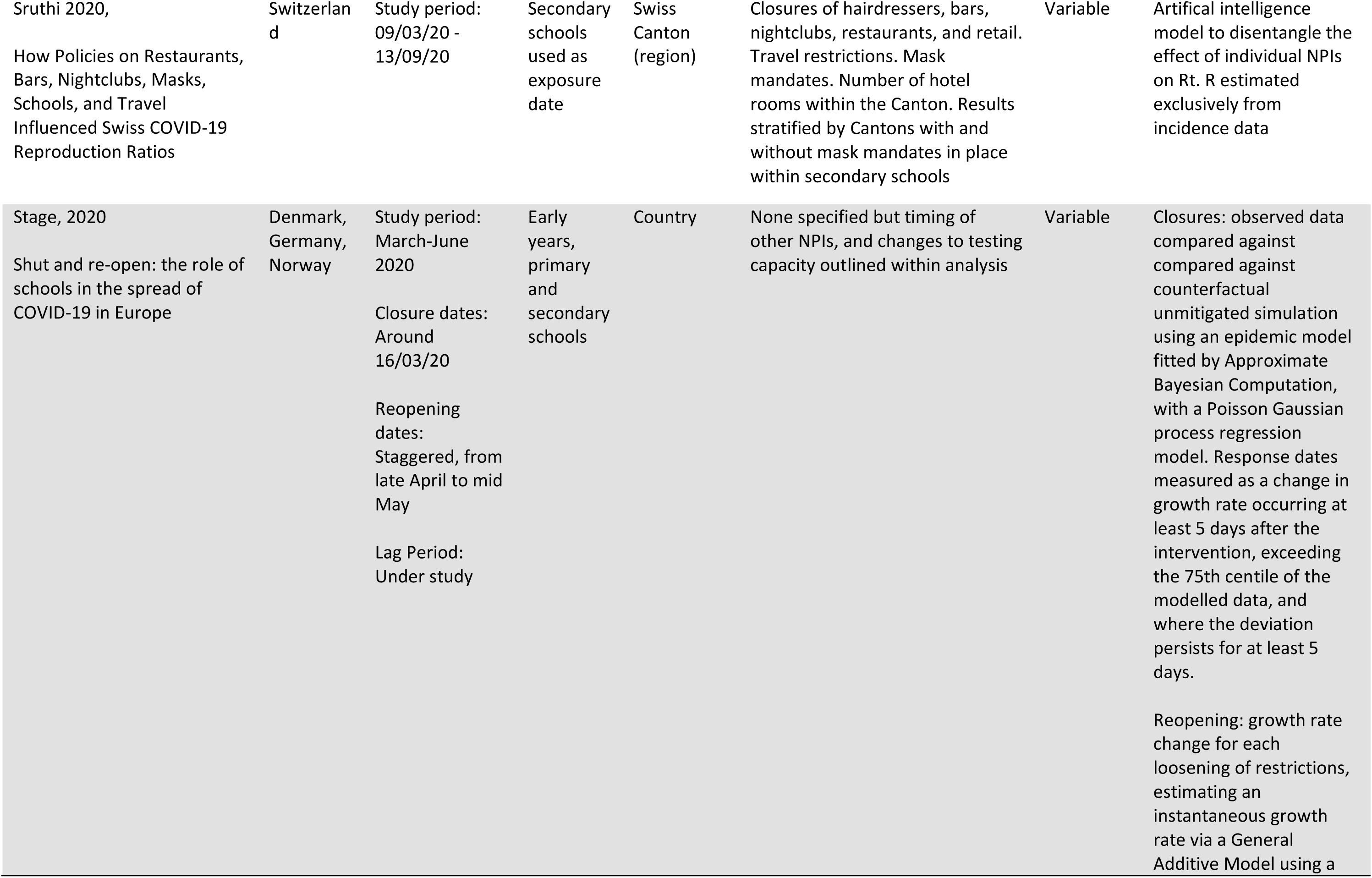
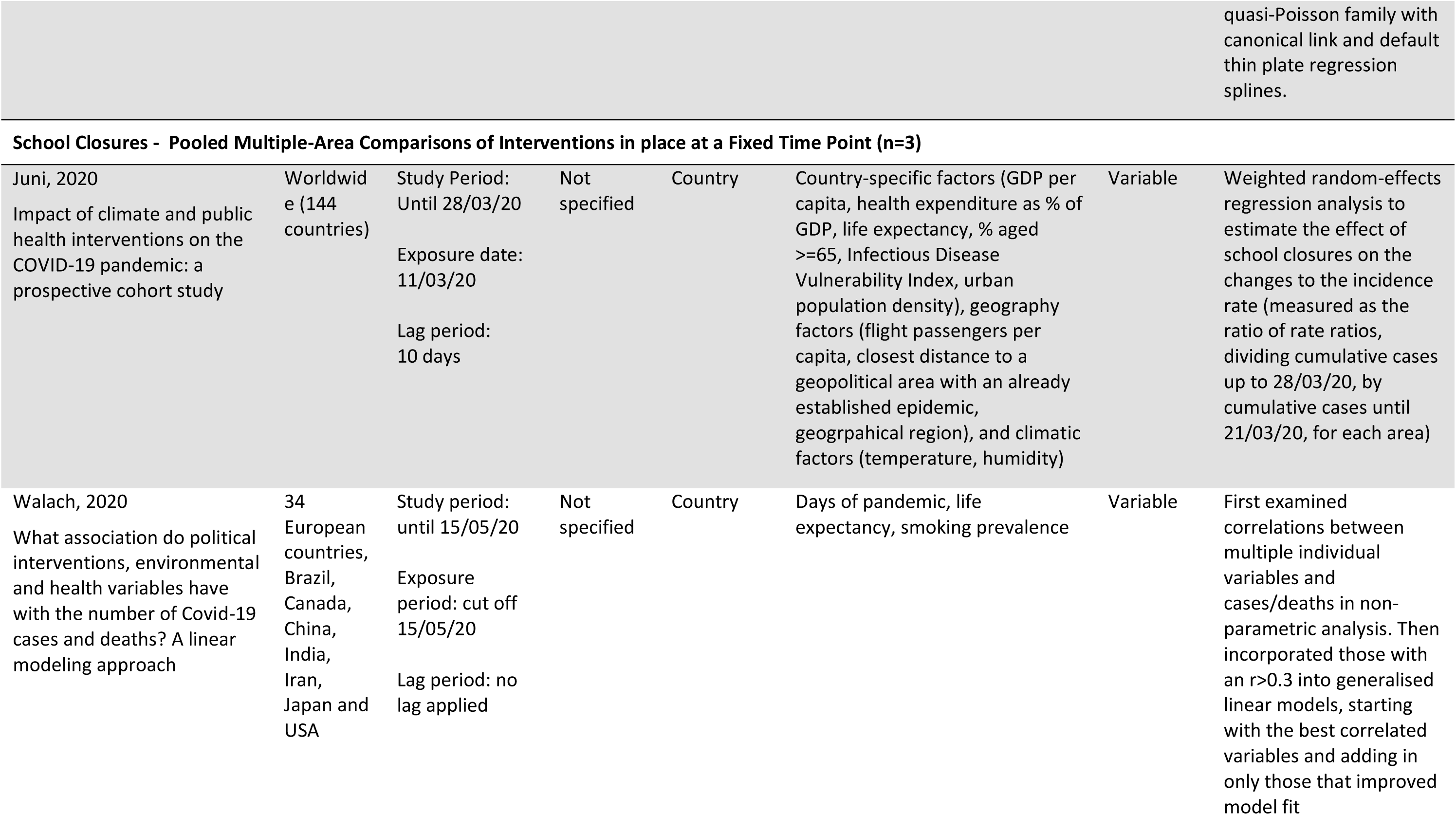
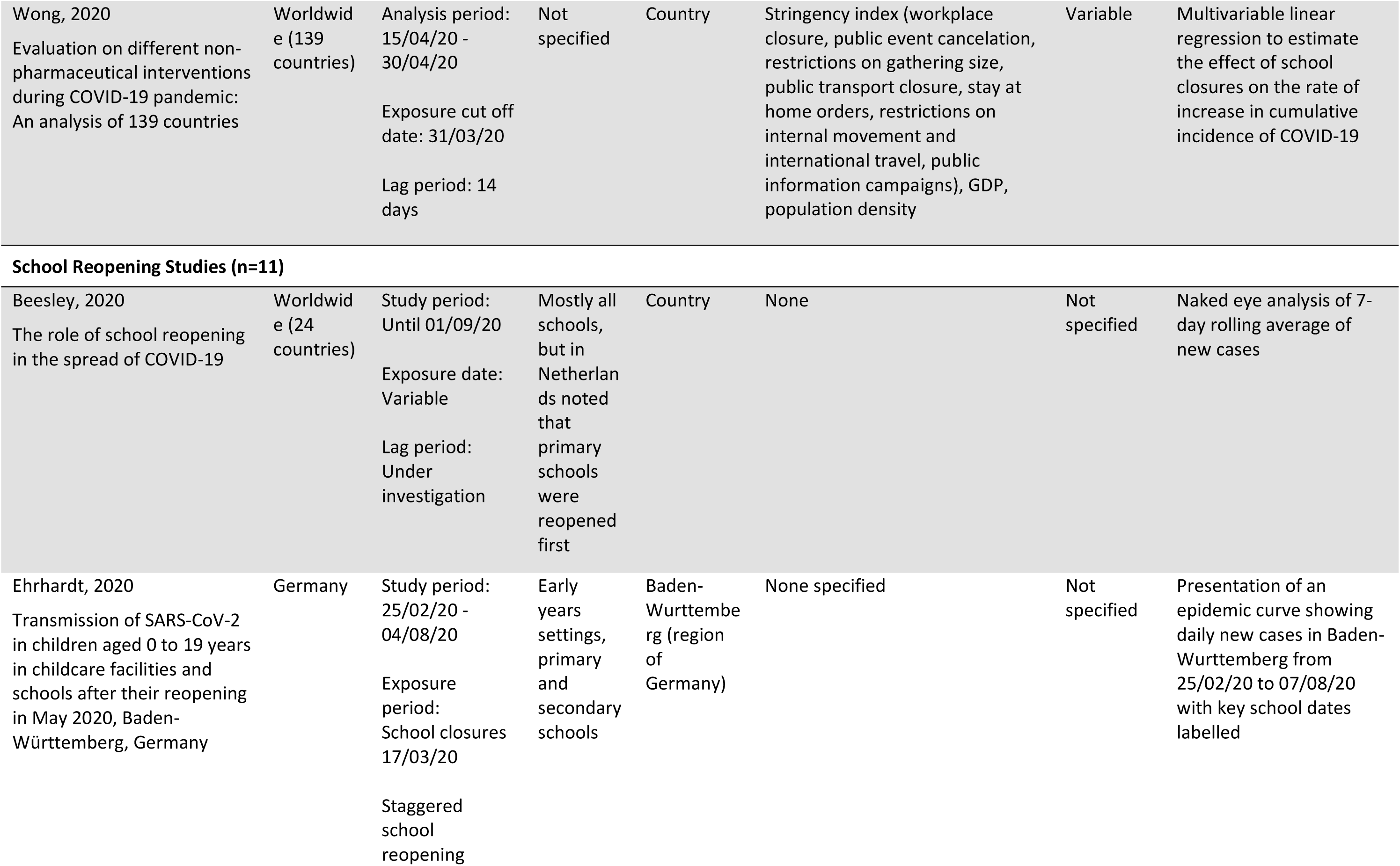
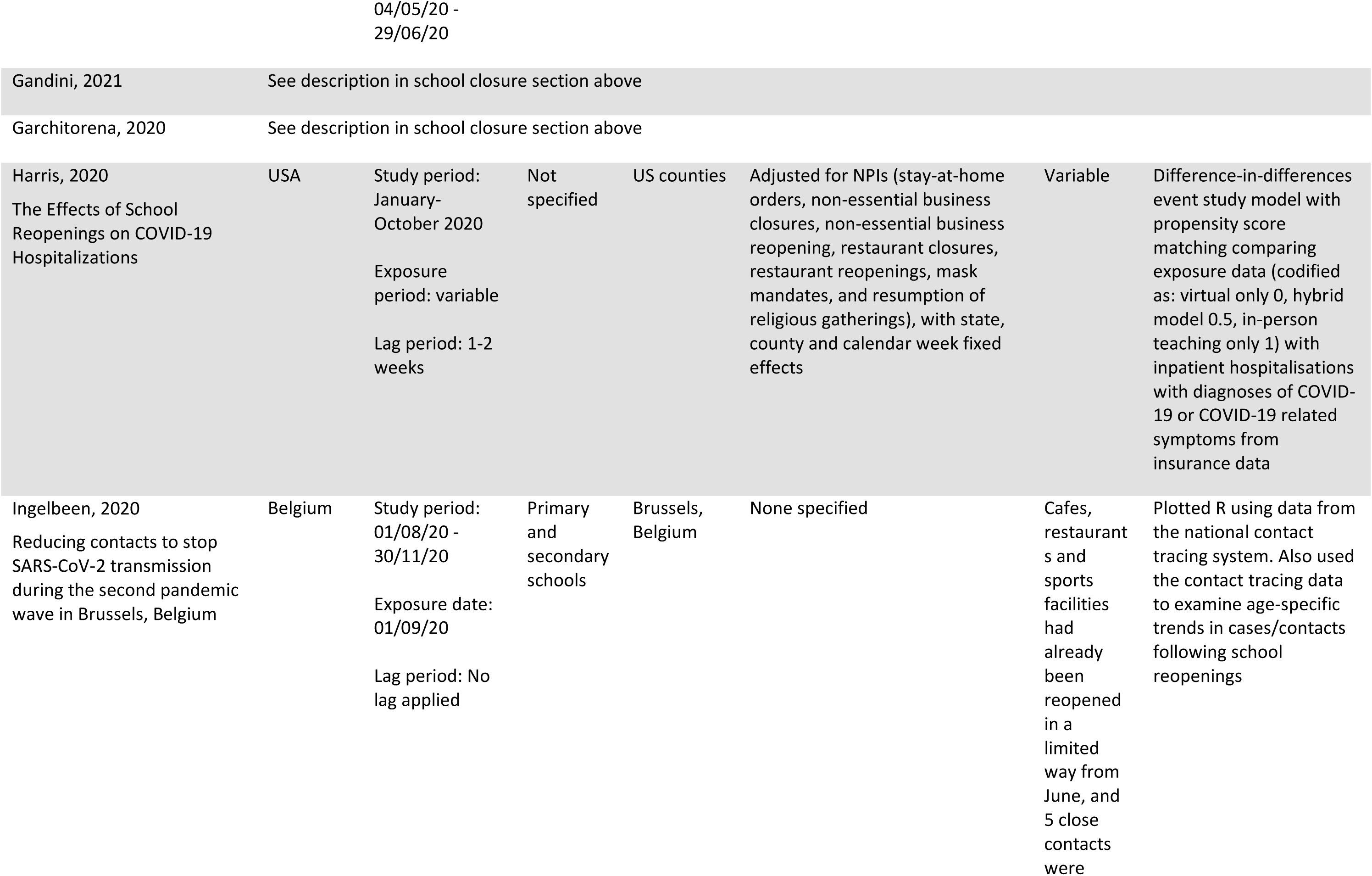
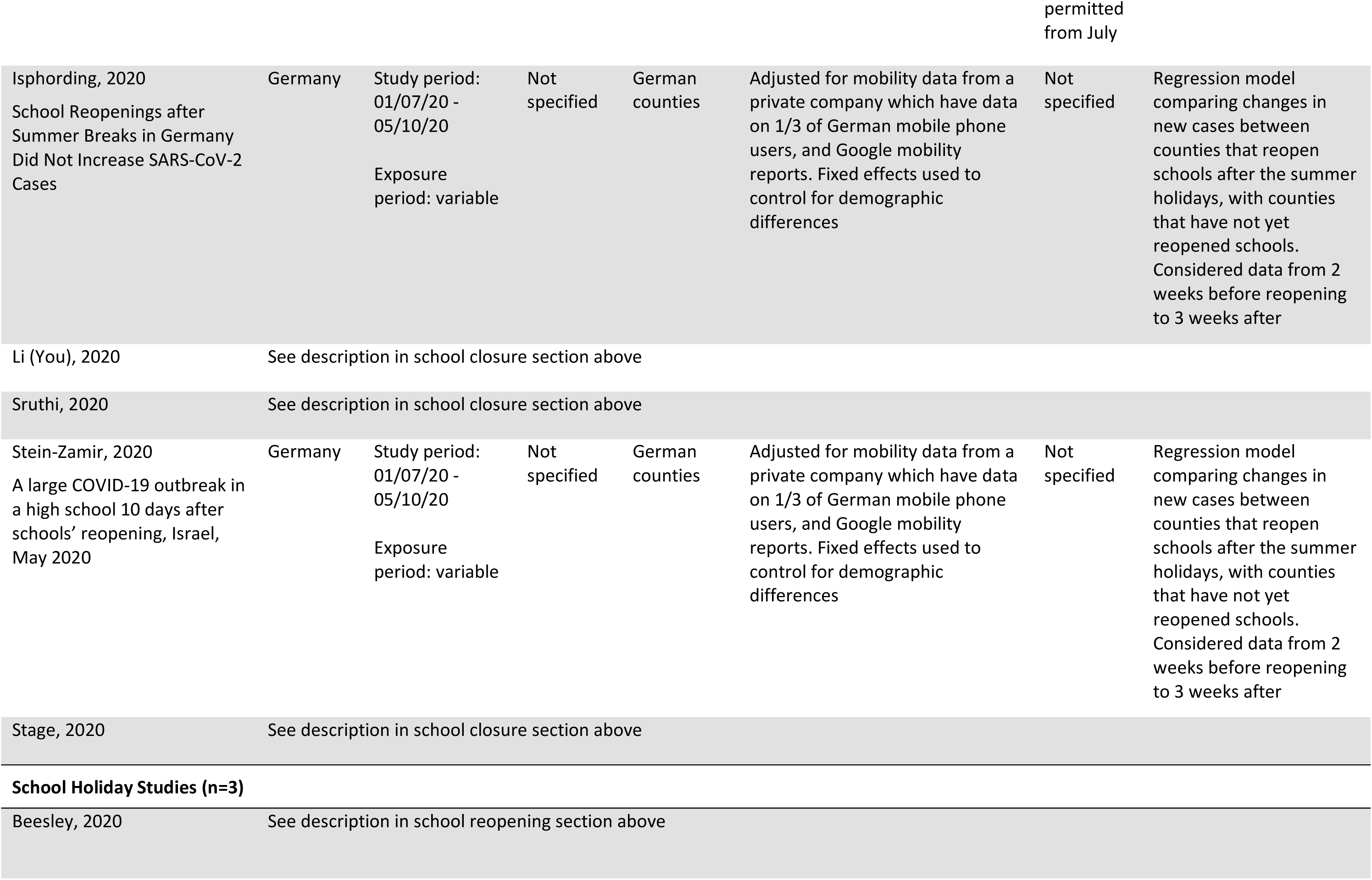
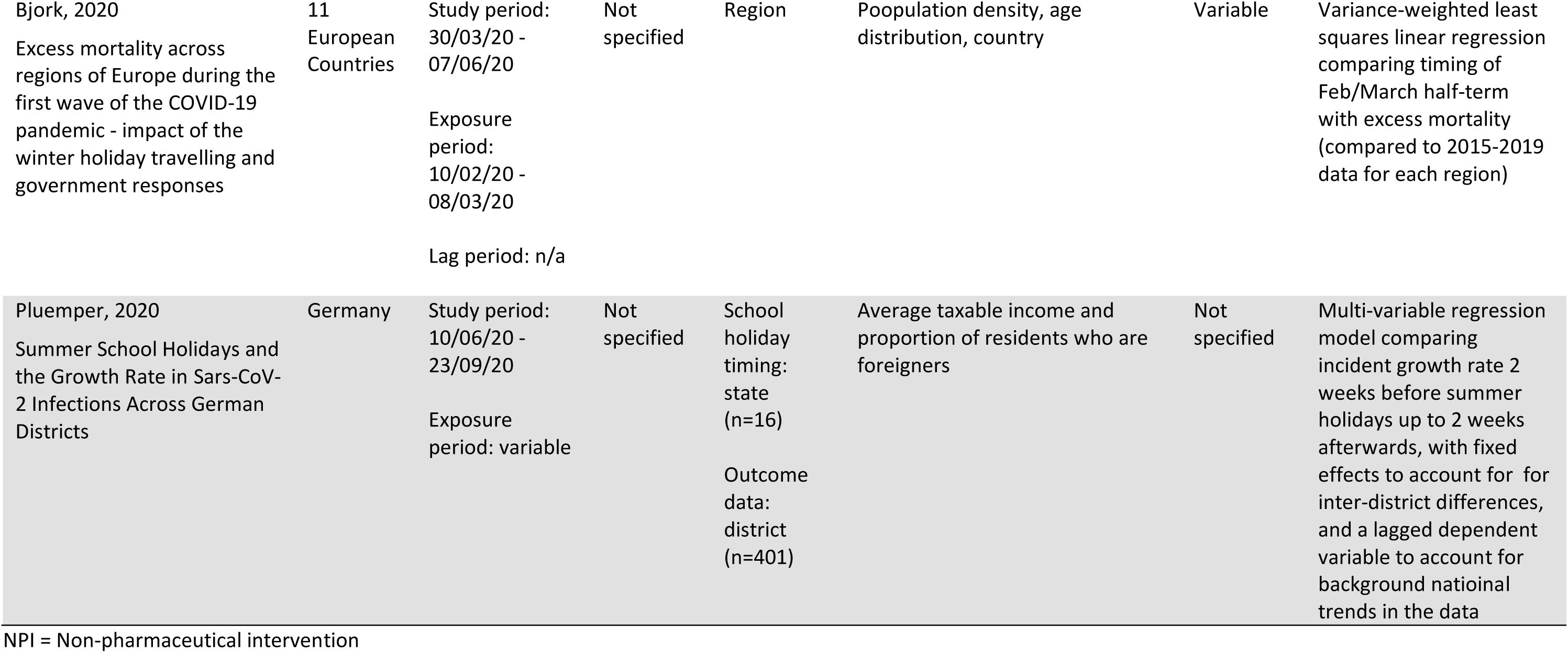
Characteristics of included studies, stratified by study design.

All studies were ecological in nature, i.e. the unit of analysis was national or regional. Of the school closure studies, 13 reported data from a single country or region (the USA (n=10)(14,19–21,33,37,42,47–49), Italy (n=1)(23), Japan (n=1)(29), and Switzerland (n=1)(43)); four reported discrete estimates for several countries(26,38,44,53); and 15 studies pooled data from multiple countries (globally (n=8)(31,34– 36,39,46,50,51), Europe only (n=2)(24, 30), Europe and other high income countries (n=5)(15,18,32,40,52)). The studies on school reopening generally reported on single countries (Germany (n=2)(22, 28), USA (n=1)(25), Switzerland (n=1)(43), Belgium (n=1)(27), Israel (n=1)(45), Italy (n=1)(23)), but one reported discrete estimates for three countries (Denamrk, Germany and Norway)(44), two pooled data from multiple countries globally(16, 35), and one pooled data from multiple European countries(24). Of the three school holiday studies, one reported on Germany(41), one pooled data from 24 countries globally(16), and one pooled data from multiple European countries(17).

The majority of studies (n=24) did not specify the type of school setting being studied. However, eight studies specified that they were reporting on primary and secondary schools only(14,16,18,19,27,29,37,49), and six additionally include early years settings(22–24,44,45,48). The two remaining studies used the date of primary school (n=1)(15) or secondary school (n=1)(43) closure as their exposure date, but did not indicate this was temporally distinct from closure of the other setting. Very few studies reported independent effect sizes for different setting types: two closure studies(24, 48) and four reopening studies(16,22,24,44).

Studies that specifically sought to estimate an effect of school closure policy on SARS-CoV-2 transmission included eight school closure studies(14,23,29,32,37,38,42,44), six school reopening studies(22,23,25,28,44,45), and three school holiday studies. The remaining studies primarily sought to estimate the effect of NPIs (but reported an independent estimate for schools, alongside estimates for other NPIs within their analysis).

The studies utilised different analytic approaches: regression models (n=24)(14,17,19– 21,25,26,28,30,31,33,35,36,39–42,44,46,48,49,51–53), Bayesian modelling (n=3)(15,18,47), comparison to a synthetic control group (n=4)(24,34,38,44), machine learning approaches (n=2)(43, 50), time series analysis (n=1)(29), and visual representation of changes in transmission over time compared against the timing of school policy interventions, with or without formal statistical analysis (n=4)(16,22,37,45). We identified three study designs used to estimate the effect of school closures: pooled multiple-area before-after comparisons (n=22)(14,15,18–21,24,26,30,32–36,39,40,42,46–50), within-area before-after comparisons(n=7)(23,29,37,38,43,44,53), and pooled multiple-area comparisons of interventions in place at a fixed time point (n=3)(31,51,52).

In most instances of school closures, particularly in European countries, other NPIs were introduced at or around the same time. Some studies dealt with this at the design stage, choosing to study places where school closures were done in (relative) isolation(37) and some at the analytical stage (typically by undertaking regression and having multiple comparator countries). Some studies did not appear to have a mechanism in place to deal with this potential confounding(32,40,44,52). Studies which pooled data from multiple areas also adjusted for other potential confounders, such as population factors (e.g. proportion of population aged ≥65, population density) and SARS-CoV-2 testing regimes.

Among school closure studies, 18 studies(14,15,19,20,24,26,29,31–34,37,39,42–44,50,51) reported effects on incidence, 11(14,19,21,30,38–40,42,46,52,53) on mortality, one(37) on hospital admissioins and mortality, and eight(18,21,23,35,36,43,47,48) on an estimate of the effective Reproductive number (R) (derived from incidence and/or mortality data). Of the school reopening studies, six reported effects on incidence(16,22,24,28,44,45), two on hospitalisations(25, 44), and four on R(23,27,35,43). Two school holiday studies reported an effect on incidence(16, 41), while the other reported on mortality(17). The assumed lag period from school policy changes to changes in incidence rate varied between seven and 20 days, with longer time periods of 26 to 28 days generally assumed for mortality.

Risk of bias is summarised in Table 2. Of the school closure studies, 14 were found to be at moderate risk of bias(14,15,18–20,24,26,30,35–37,46–48), 14 at serious risk(21,23,29,31,33,34,38,39,42,43,49–51,53), and four at critical risk of bias(32,40,44,52). Of the school reopening studies, four were found to be at moderate risk(24,25,28,35), four at serious risk(23,27,43,44), and three at critical risk of bias(16,22,45). The school holiday studies were found to be at moderate (n=1)(41), serious (n=1)(17), or critical (n=1)(16) risk of bias.

**Table 2:**
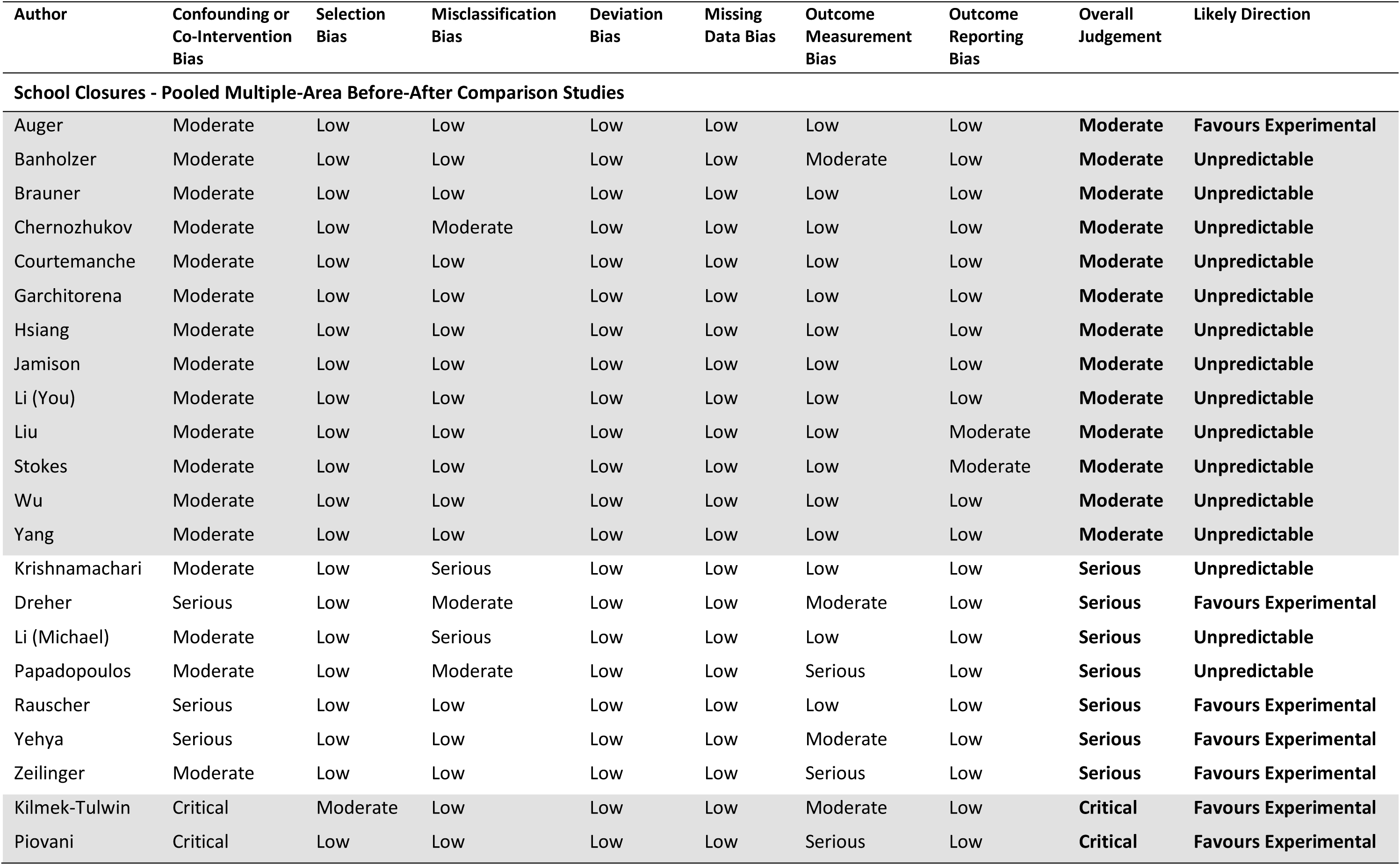
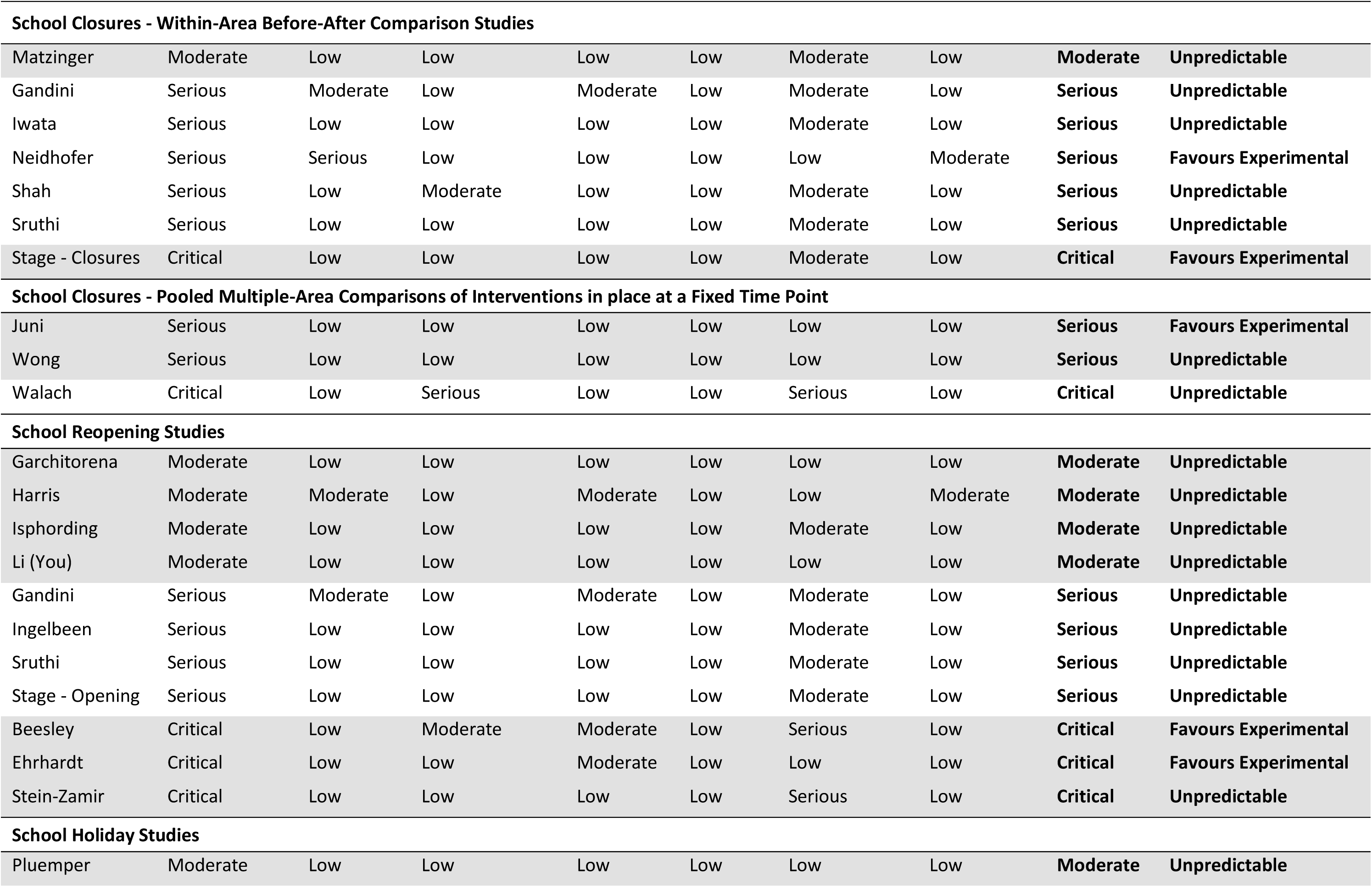
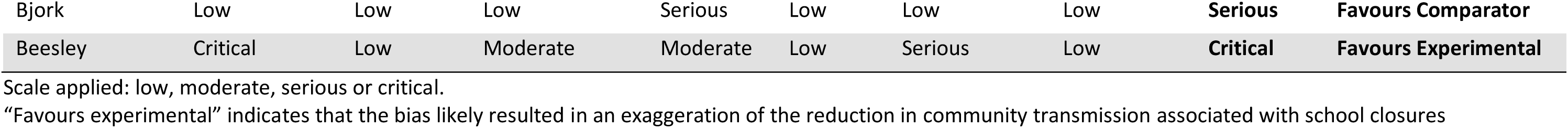
Findings from the risk of bias assessment using the ROBINS-I tool, stratified by study design.

| Author | Confounding or Co-Intervention Bias | Selection Bias | Misclassification Bias | Deviation Bias | Missing Data Bias | Outcome Measurement Bias | Outcome Reporting Bias | Overall Judgement | Likely Direction |
| --- | --- | --- | --- | --- | --- | --- | --- | --- | --- |
| **School Closures - Pooled Multiple-Area Before-After Comparison Studies** |  |  |  |  |  |  |  |  |  |
| Auger | Moderate | Low | Low | Low | Low | Low | Low | **Moderate** | **Favours Experimental** |
| Banholzer | Moderate | Low | Low | Low | Low | Moderate | Low | **Moderate** | **Unpredictable** |
| Brauner | Moderate | Low | Low | Low | Low | Low | Low | **Moderate** | **Unpredictable** |
| Chernozhukov | Moderate | Low | Moderate | Low | Low | Low | Low | **Moderate** | **Unpredictable** |
| Courtemanche | Moderate | Low | Low | Low | Low | Low | Low | **Moderate** | **Unpredictable** |
| Garchitorena | Moderate | Low | Low | Low | Low | Low | Low | **Moderate** | **Unpredictable** |
| Hsiang | Moderate | Low | Low | Low | Low | Low | Low | **Moderate** | **Unpredictable** |
| Jamison | Moderate | Low | Low | Low | Low | Low | Low | **Moderate** | **Unpredictable** |
| Li (You) | Moderate | Low | Low | Low | Low | Low | Low | **Moderate** | **Unpredictable** |
| Liu | Moderate | Low | Low | Low | Low | Low | Moderate | **Moderate** | **Unpredictable** |
| Stokes | Moderate | Low | Low | Low | Low | Low | Moderate | **Moderate** | **Unpredictable** |
| Wu | Moderate | Low | Low | Low | Low | Low | Low | **Moderate** | **Unpredictable** |
| Yang | Moderate | Low | Low | Low | Low | Low | Low | **Moderate** | **Unpredictable** |
| Krishnamachari | Moderate | Low | Serious | Low | Low | Low | Low | **Serious** | **Unpredictable** |
| Dreher | Serious | Low | Moderate | Low | Low | Moderate | Low | **Serious** | **Favours Experimental** |
| Li (Michael) | Moderate | Low | Serious | Low | Low | Low | Low | **Serious** | **Unpredictable** |
| Papadopoulos | Moderate | Low | Moderate | Low | Low | Serious | Low | **Serious** | **Unpredictable** |
| Rauscher | Serious | Low | Low | Low | Low | Low | Low | **Serious** | **Favours Experimental** |
| Yehya | Serious | Low | Low | Low | Low | Moderate | Low | **Serious** | **Favours Experimental** |
| Zeilinger | Moderate | Low | Low | Low | Low | Serious | Low | **Serious** | **Favours Experimental** |
| Kilmek-Tulwin | Critical | Moderate | Low | Low | Low | Moderate | Low | **Critical** | **Favours Experimental** |
| Piovani | Critical | Low | Low | Low | Low | Serious | Low | **Critical** | **Favours Experimental** |

| **School Closures - Within-Area Before-After Comparison Studies** |  |  |  |  |  |  |  |  |  |
| --- | --- | --- | --- | --- | --- | --- | --- | --- | --- |
| Matzinger | Moderate | Low | Low | Low | Low | Moderate | Low | **Moderate** | **Unpredictable** |
| Gandini | Serious | Moderate | Low | Moderate | Low | Moderate | Low | **Serious** | **Unpredictable** |
| Iwata | Serious | Low | Low | Low | Low | Moderate | Low | **Serious** | **Unpredictable** |
| Neidhofer | Serious | Serious | Low | Low | Low | Low | Moderate | **Serious** | **Favours Experimental** |
| Shah | Serious | Low | Moderate | Low | Low | Moderate | Low | **Serious** | **Unpredictable** |
| Sruthi | Serious | Low | Low | Low | Low | Moderate | Low | **Serious** | **Unpredictable** |
| Stage - Closures | Critical | Low | Low | Low | Low | Moderate | Low | **Critical** | **Favours Experimental** |
| **School Closures - Pooled Multiple-Area Comparisons of Interventions in place at a Fixed Time Point** |  |  |  |  |  |  |  |  |  |
| Juni | Serious | Low | Low | Low | Low | Low | Low | **Serious** | **Favours Experimental** |
| Wong | Serious | Low | Low | Low | Low | Low | Low | **Serious** | **Unpredictable** |
| Walach | Critical | Low | Serious | Low | Low | Serious | Low | **Critical** | **Unpredictable** |
| **School Reopening Studies** |  |  |  |  |  |  |  |  |  |
| Garchitoren | Moderate | Low | Low | Low | Low | Low | Low | **Moderate** | **Unpredictable** |
| Harris | Moderate | Moderate | Low | Moderate | Low | Low | Moderate | **Moderate** | **Unpredictable** |
| Isphording | Moderate | Low | Low | Low | Low | Moderate | Low | **Moderate** | **Unpredictable** |
| Li (You) | Moderate | Low | Low | Low | Low | Low | Low | **Moderate** | **Unpredictable** |
| Gandini | Serious | Moderate | Low | Moderate | Low | Moderate | Low | **Serious** | **Unpredictable** |
| Ingelbeen | Serious | Low | Low | Low | Low | Moderate | Low | **Serious** | **Unpredictable** |
| Sruthi | Serious | Low | Low | Low | Low | Moderate | Low | **Serious** | **Unpredictable** |
| Stage - Opening | Serious | Low | Low | Low | Low | Moderate | Low | **Serious** | **Unpredictable** |
| Beesley | Critical | Low | Moderate | Moderate | Low | Serious | Low | **Critical** | **Favours Experimental** |
| Ehrhardt | Critical | Low | Low | Moderate | Low | Low | Low | **Critical** | **Favours Experimental** |
| Stein-Zamir | Critical | Low | Low | Low | Low | Serious | Low | **Critical** | **Unpredictable** |
| **School Holiday Studies** |  |  |  |  |  |  |  |  |  |
| Pluemper | Moderate | Low | Low | Low | Low | Low | Low | **Moderate** | **Unpredictable** |
| Bjork | Low | Low | Low | Serious | Low | Low | Low | **Serious** | **Favours Comparator** |
| Beesley | Critical | Low | Moderate | Moderate | Low | Serious | Low | **Critical** | **Favours Experimental** |

Scale applied: low, moderate, serious or critical.
“Favours experimental” indicates that the bias likely resulted in an exaggeration of the reduction in community transmission associated with school closures

There was significant heterogeneity in the study findings (table 3): 17 studies(14,24,31,32,34–38,40,42–44,48–51) reported that closing schools was associated with a reduction in transmission rates; nine (15,18,20,23,26,29,30,39,47) found no association between school closures and transmission; five (19,21,33,46,53) reported mixed findings with evidence of a reduction in transmission in some analyses but not others; and one study(52) reported that school closures were associated with an increase in mortality. The reported effect size of closing schools ranged from precise estimates of no effect(26) to approximately halving the incidence(14); and from approximately doubling mortality(52), to approximately halving mortality(14). The studies at the highest risk of bias generally reported large reductions in transmission associated with school closures, while studies at lower levels of bias reported more variable findings (figure 2). Of the school reopening studies, six(22–25,28,44) reported no increase in transmission associated with reopening of schools, while two(16, 43) reported mixed findings, and three(27,35,45) reported increases in transmission. Of the four school reopening studies at lowest risk of bias(24,25,28,35), three(24,25,28) reported no association between school reopenings and transmission.

**Figure 2:**
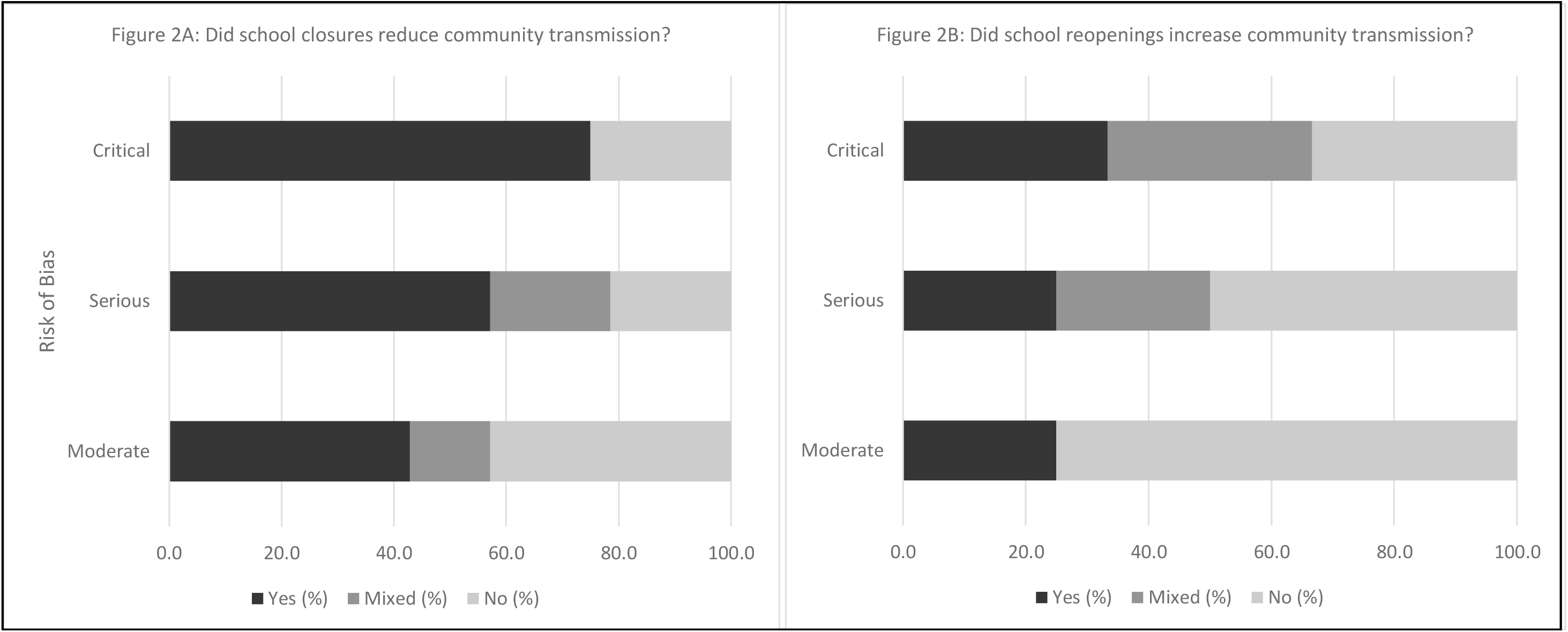
Main findings, stratified by risk of bias. Figure 2A presents the studies’ response to the question: Did school closures reduced community transmission? (Yes, No, Mixed). Figure 2B presents the studies’ response to the question: Did school reopenings increase community transmission? (Yes, No, Mixed)

**Table 3:**
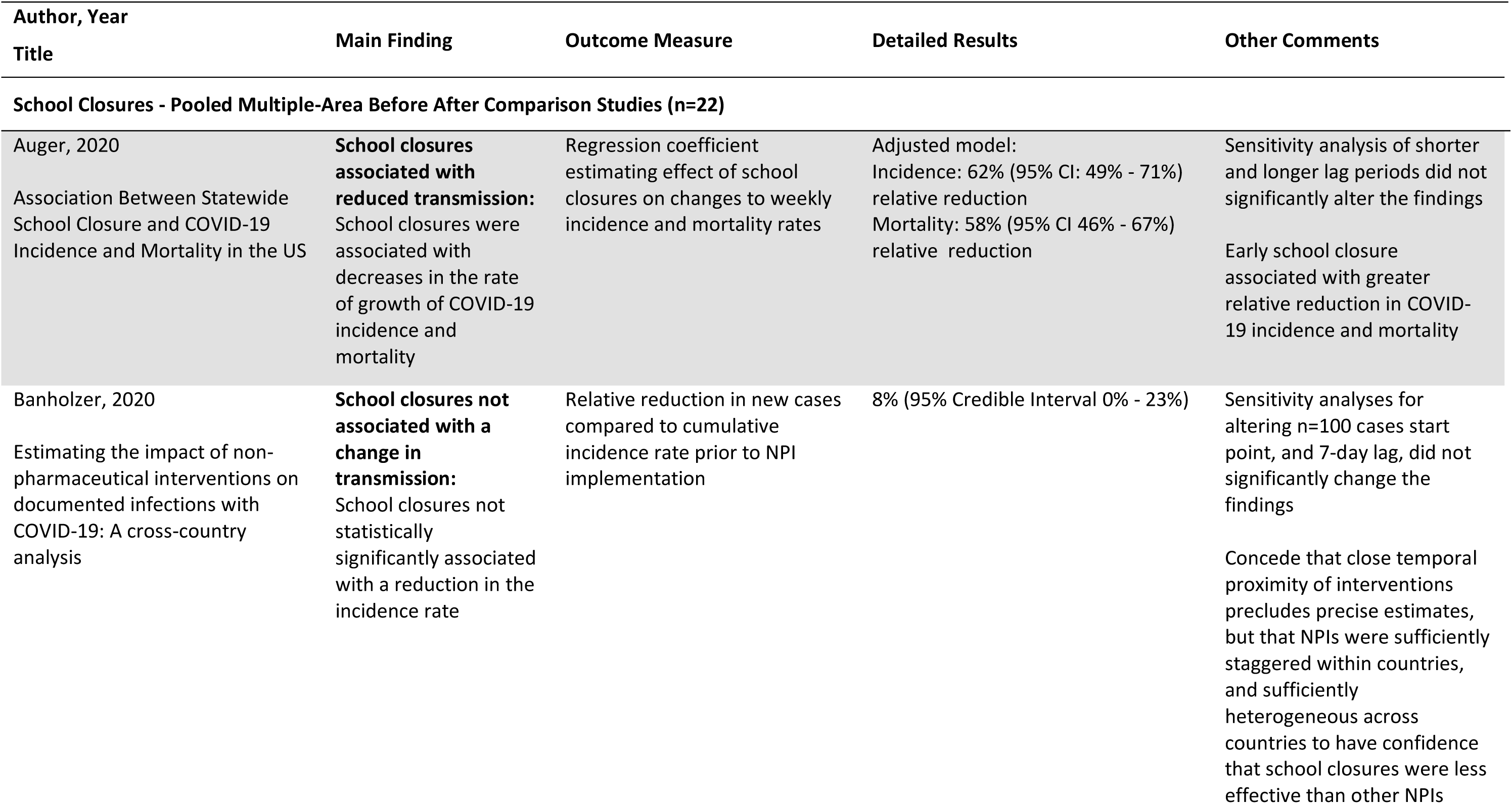
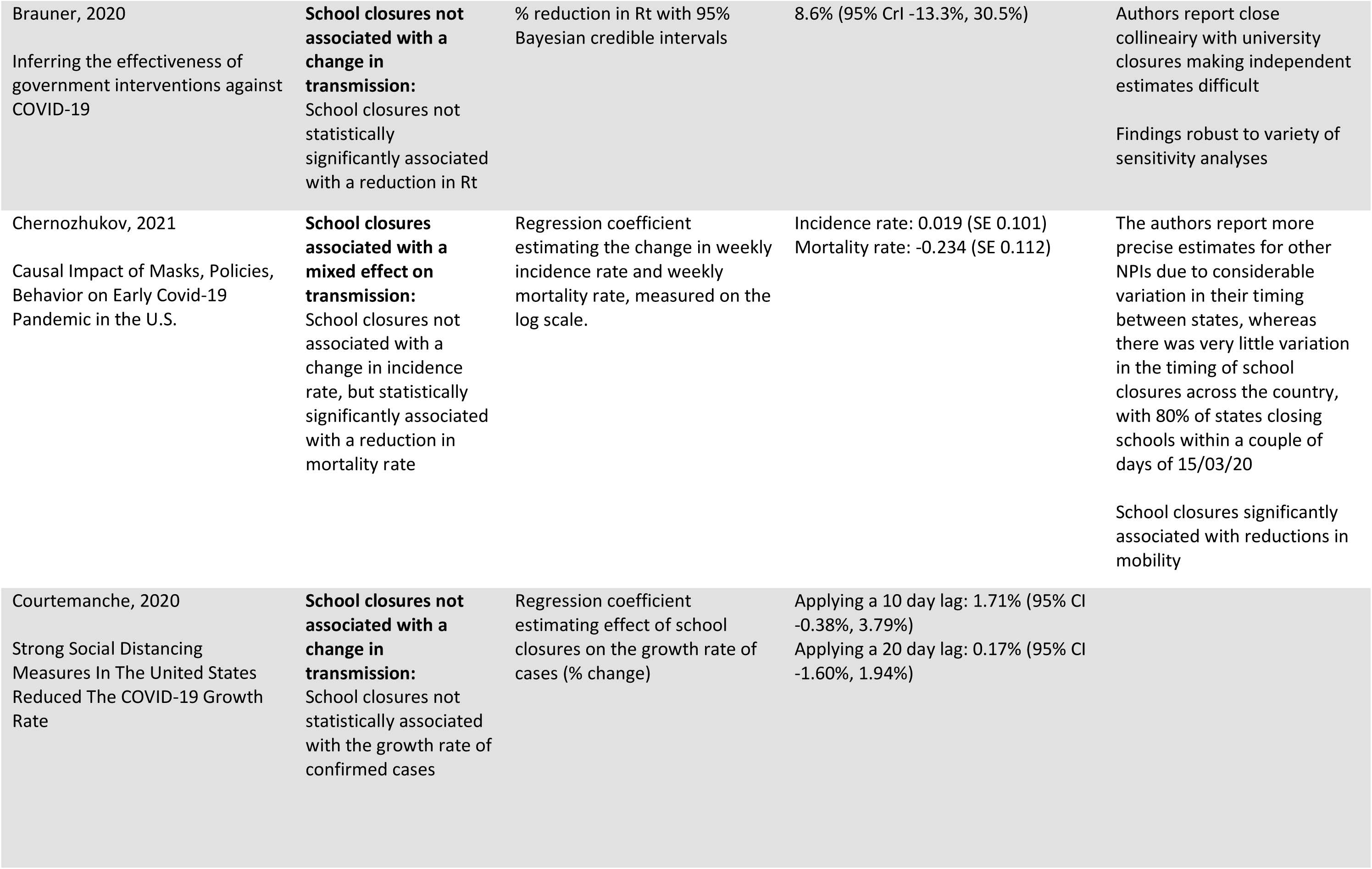
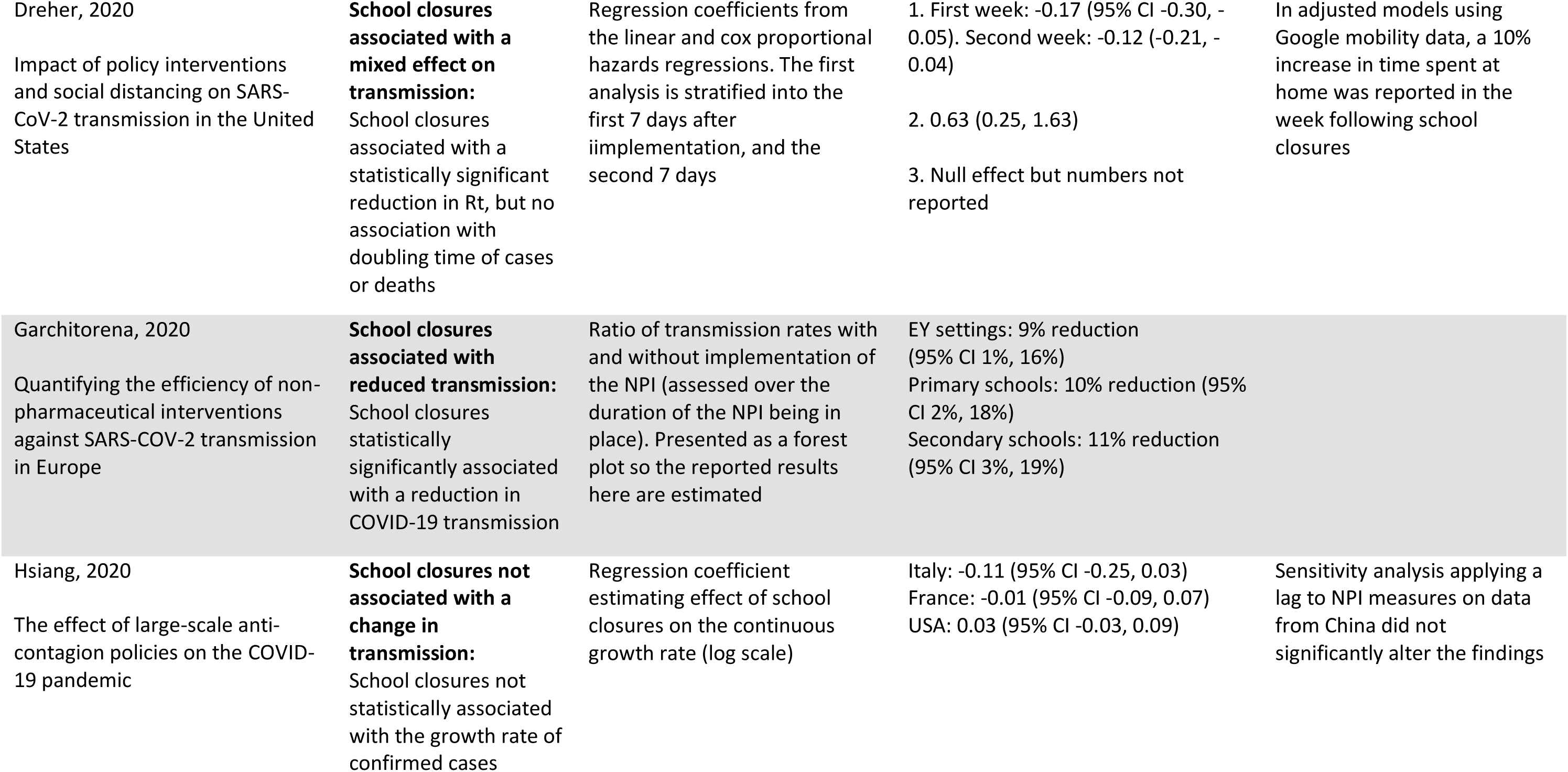
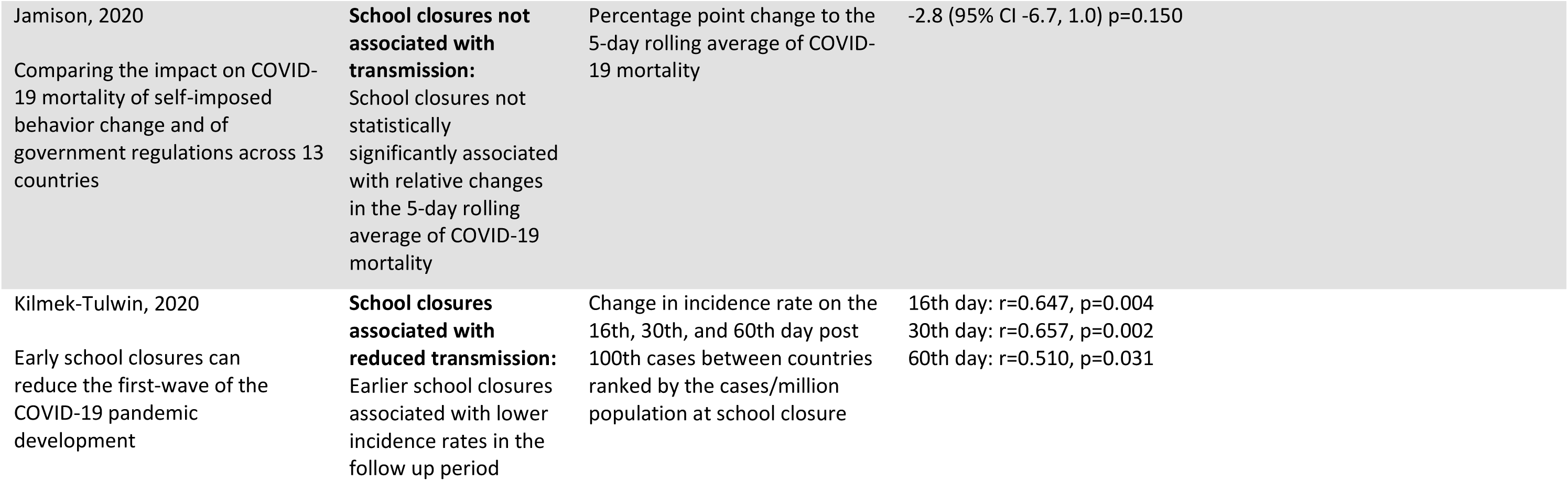
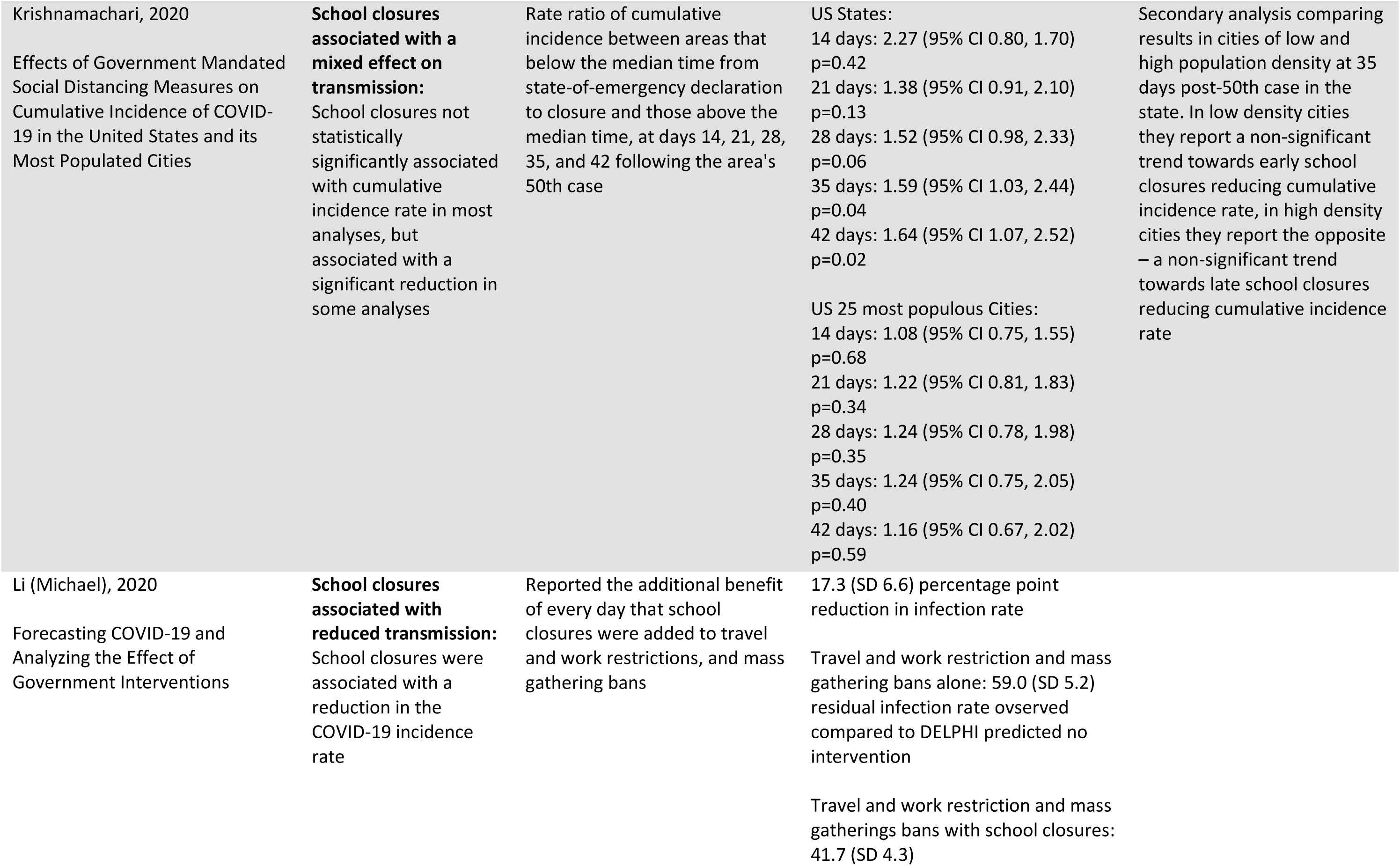
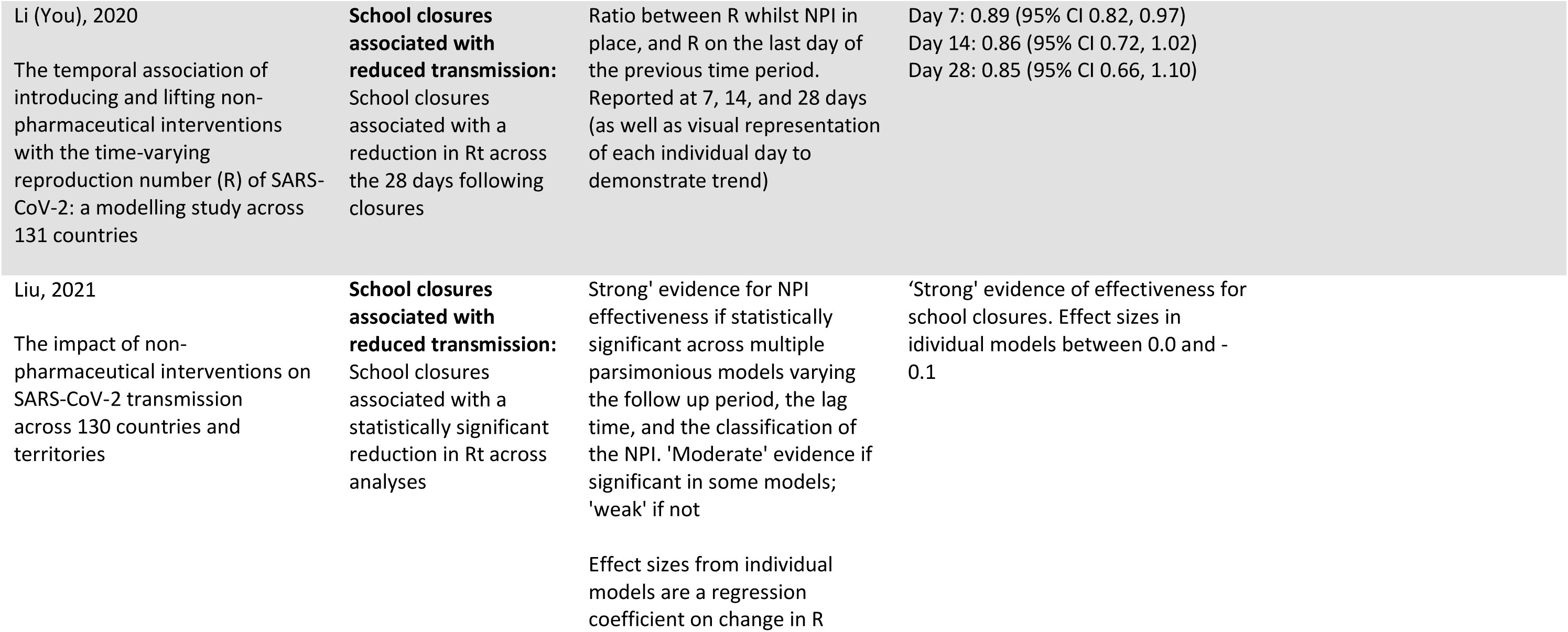
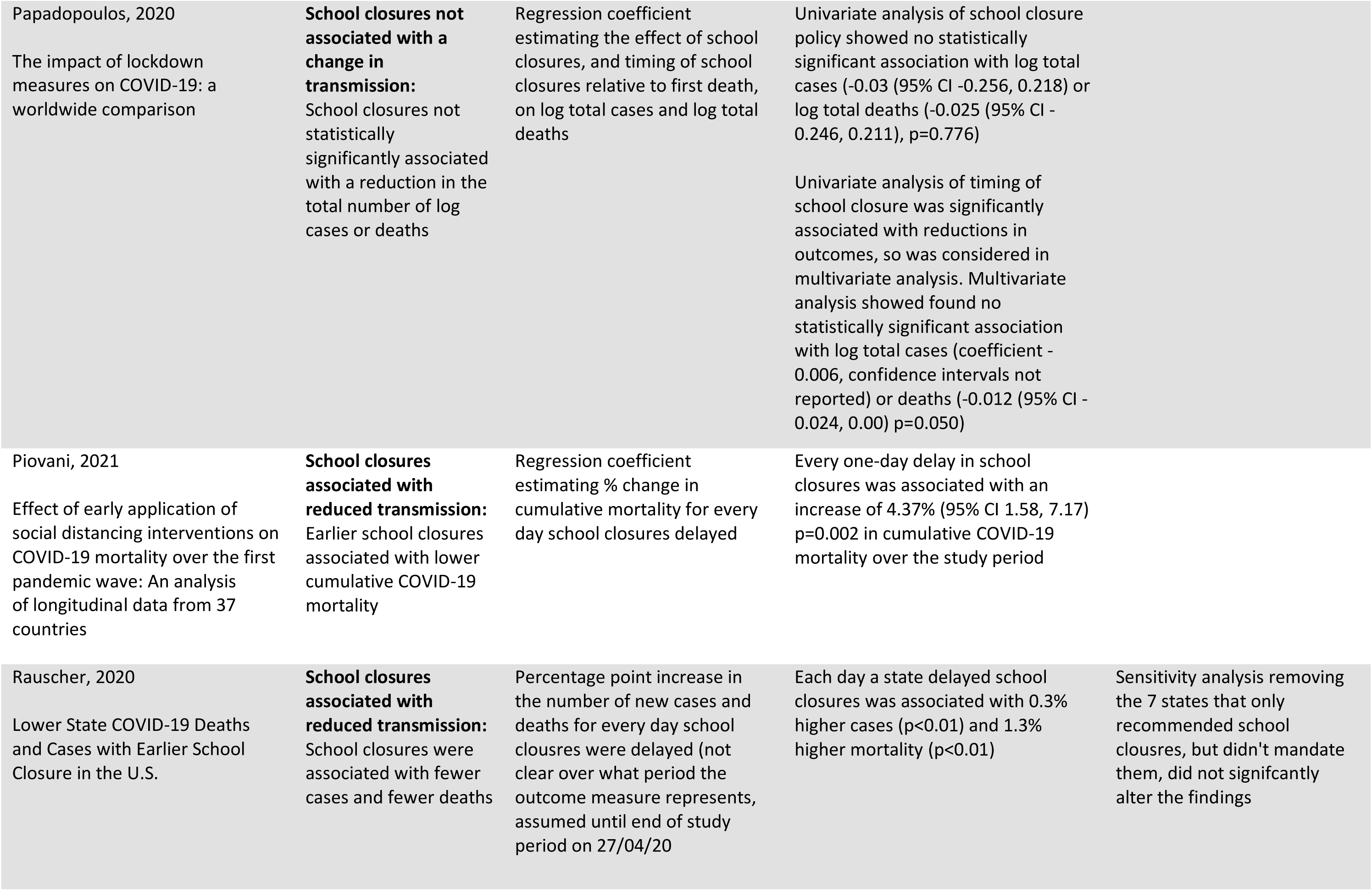
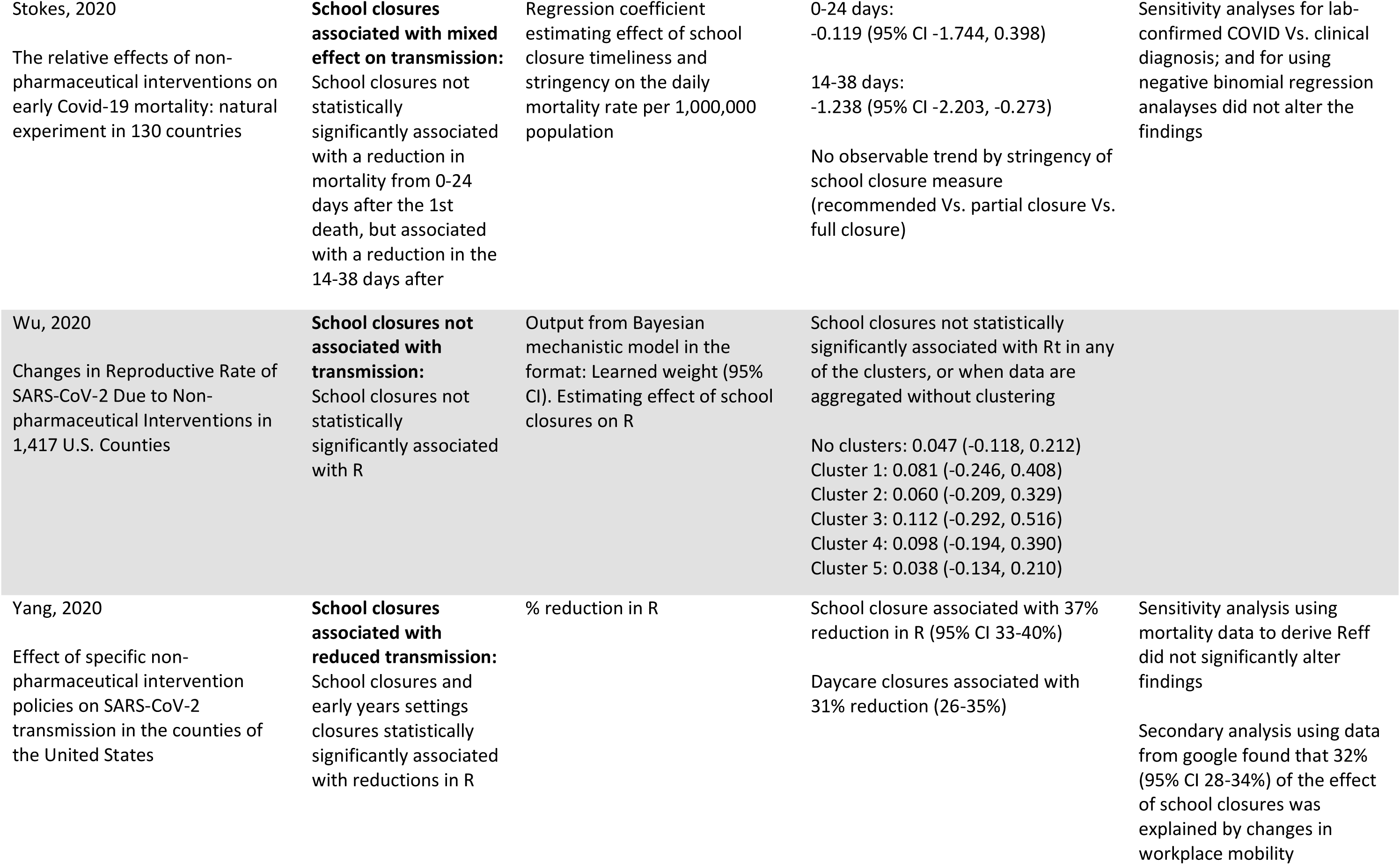
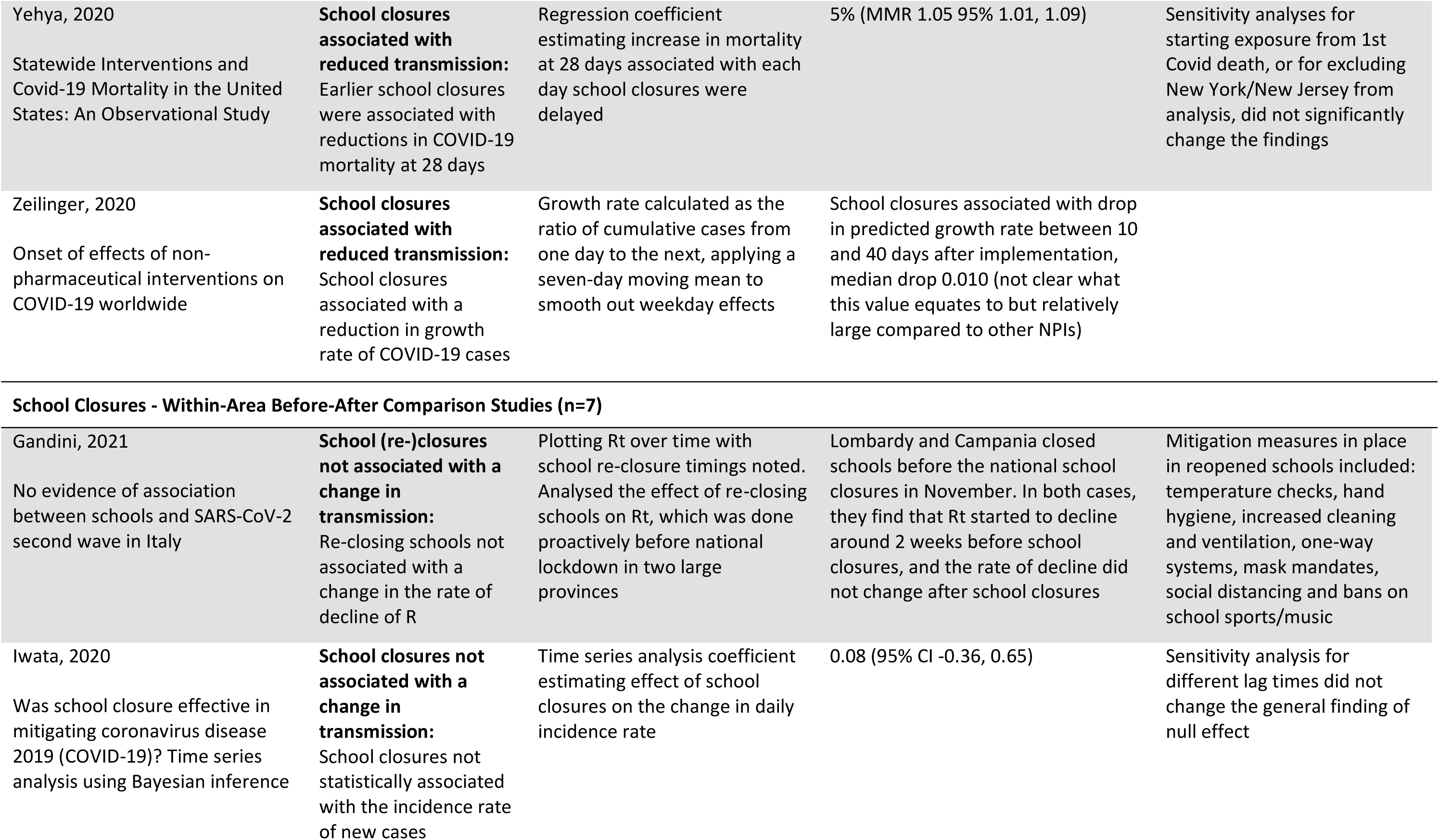
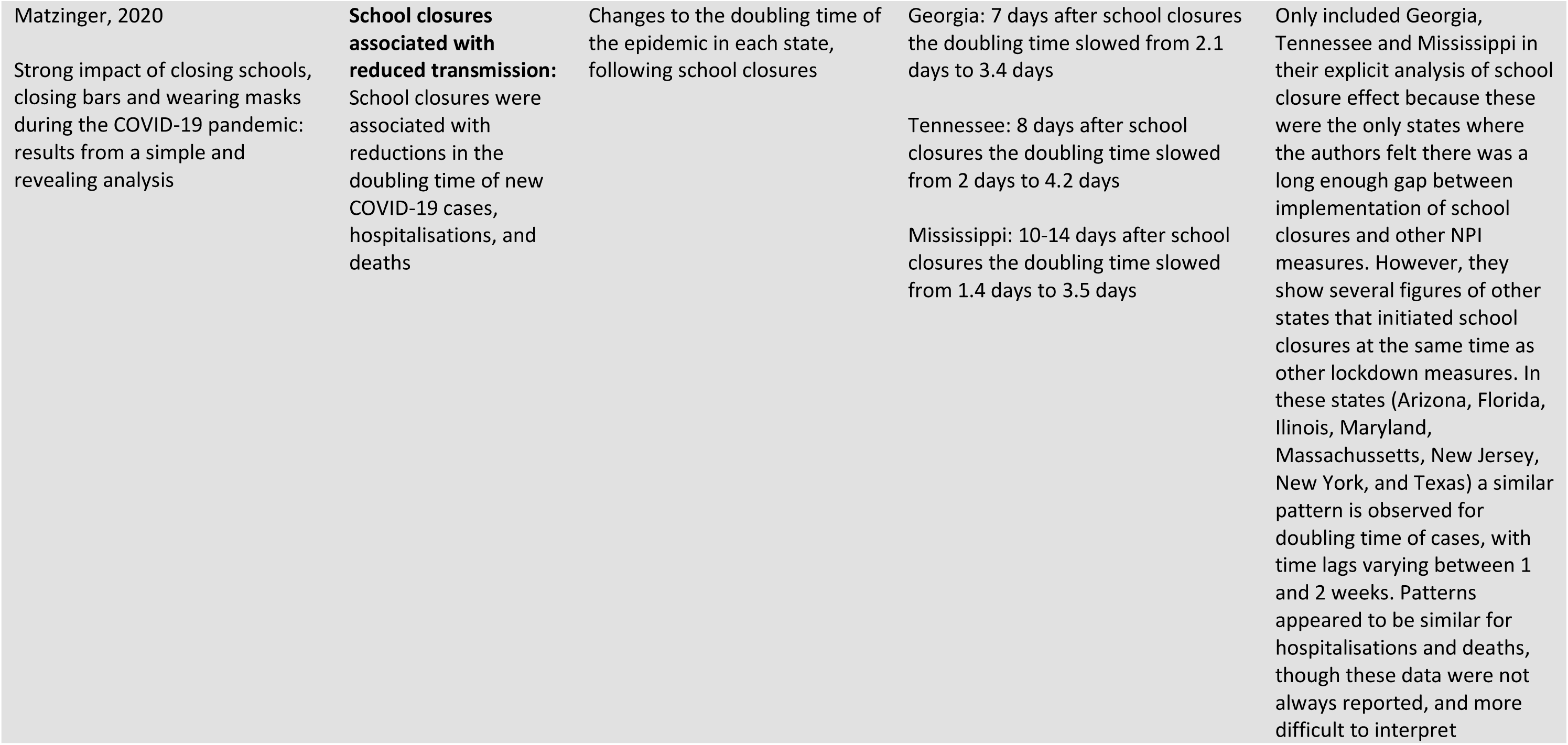
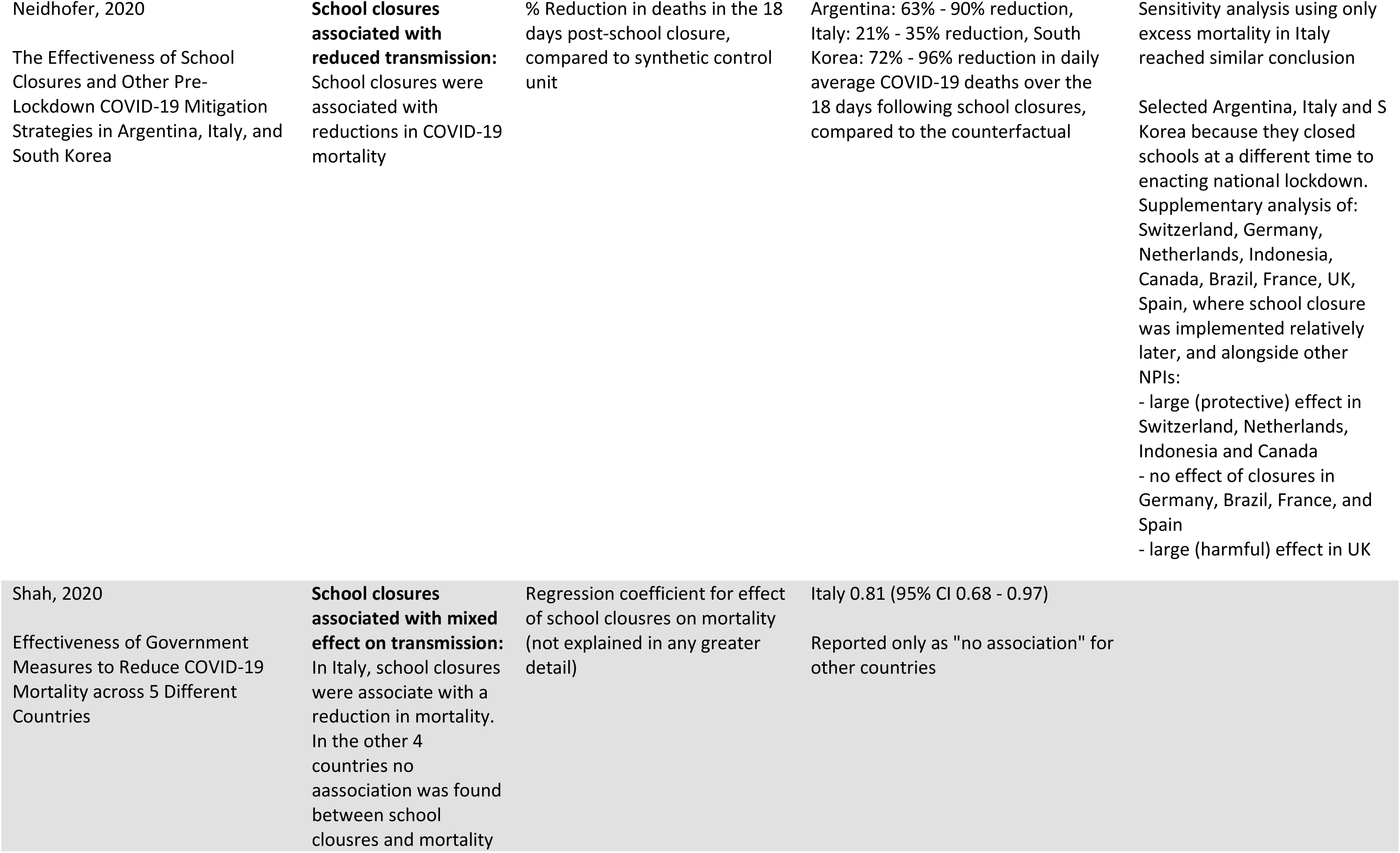
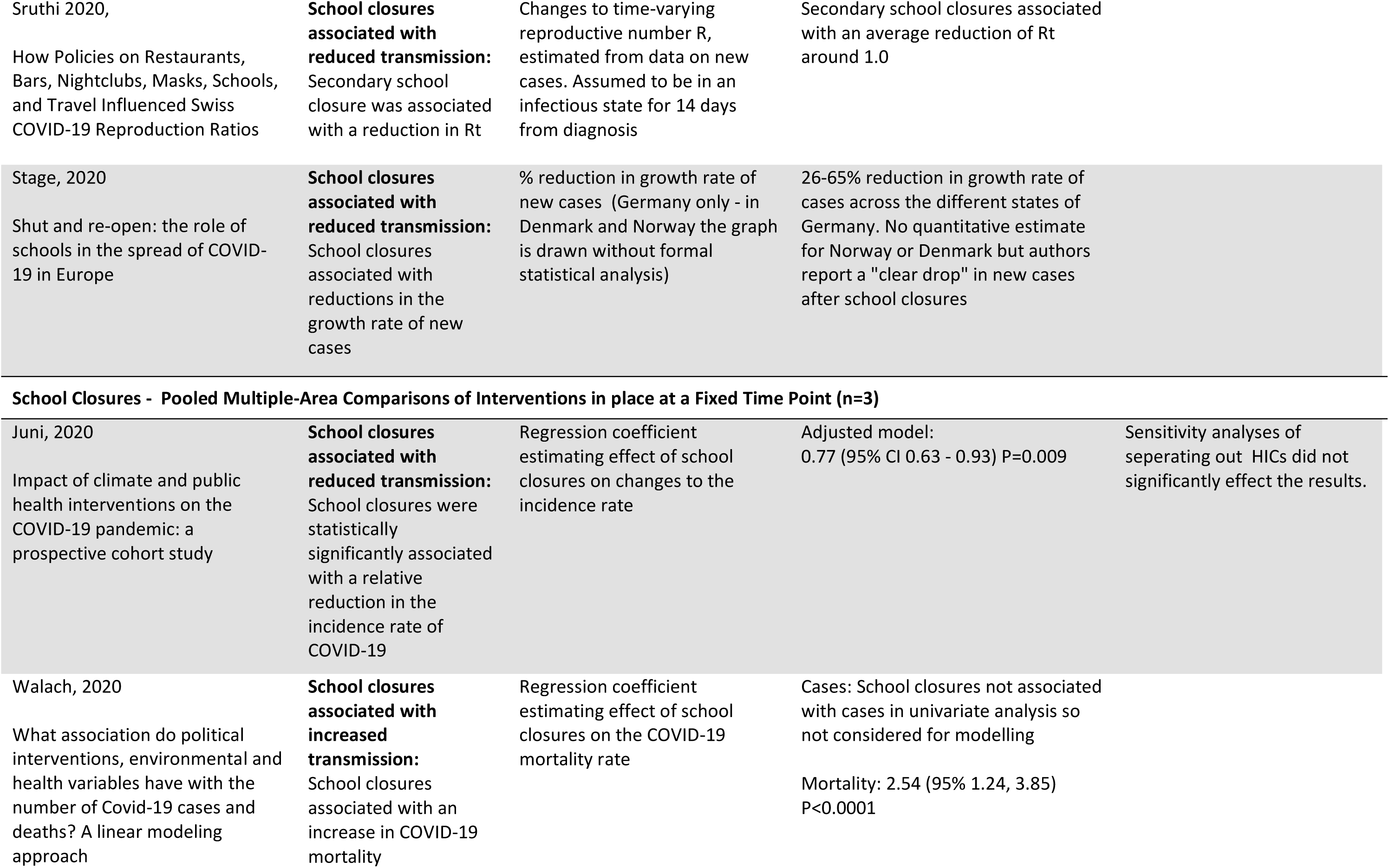
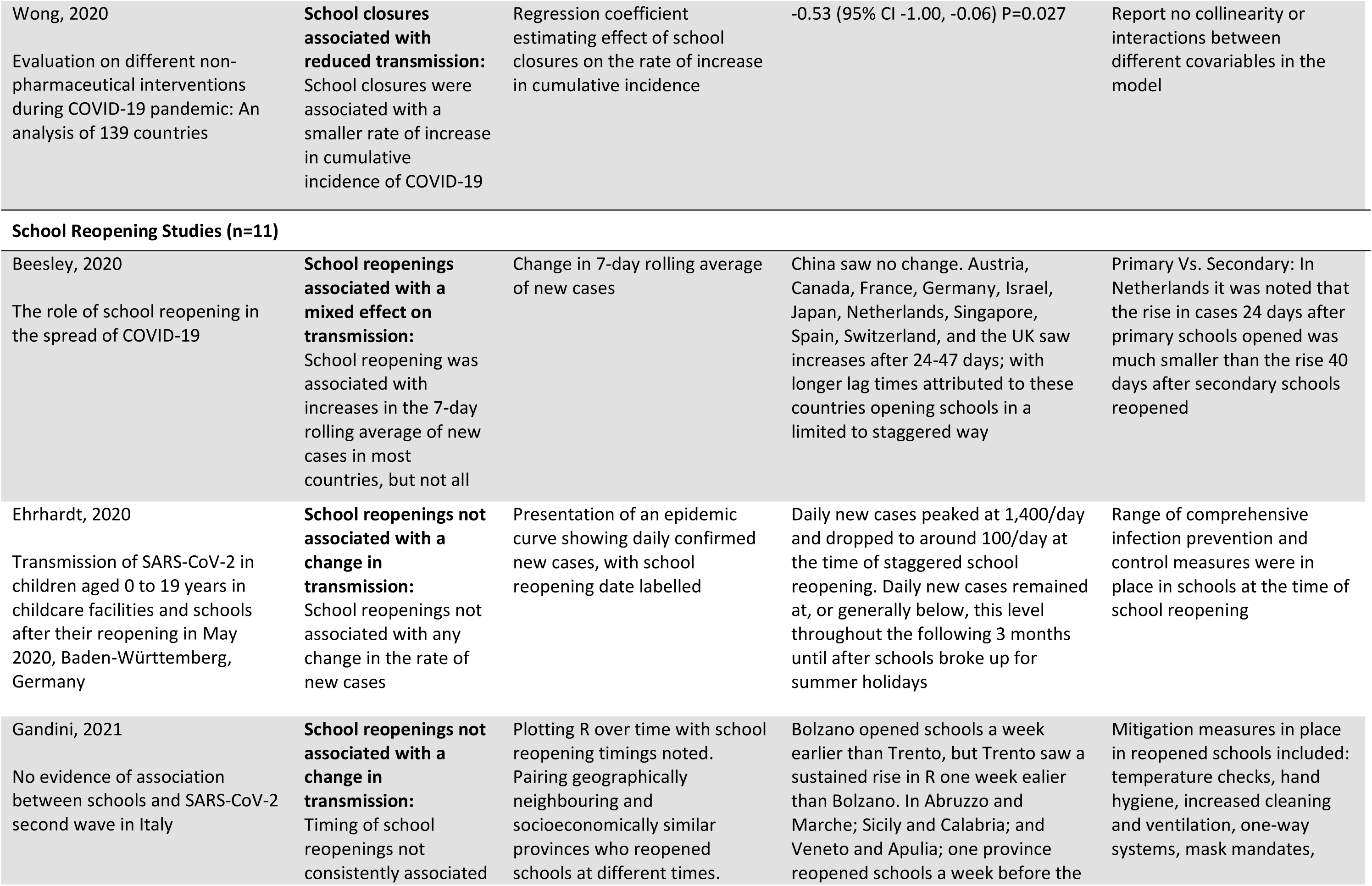
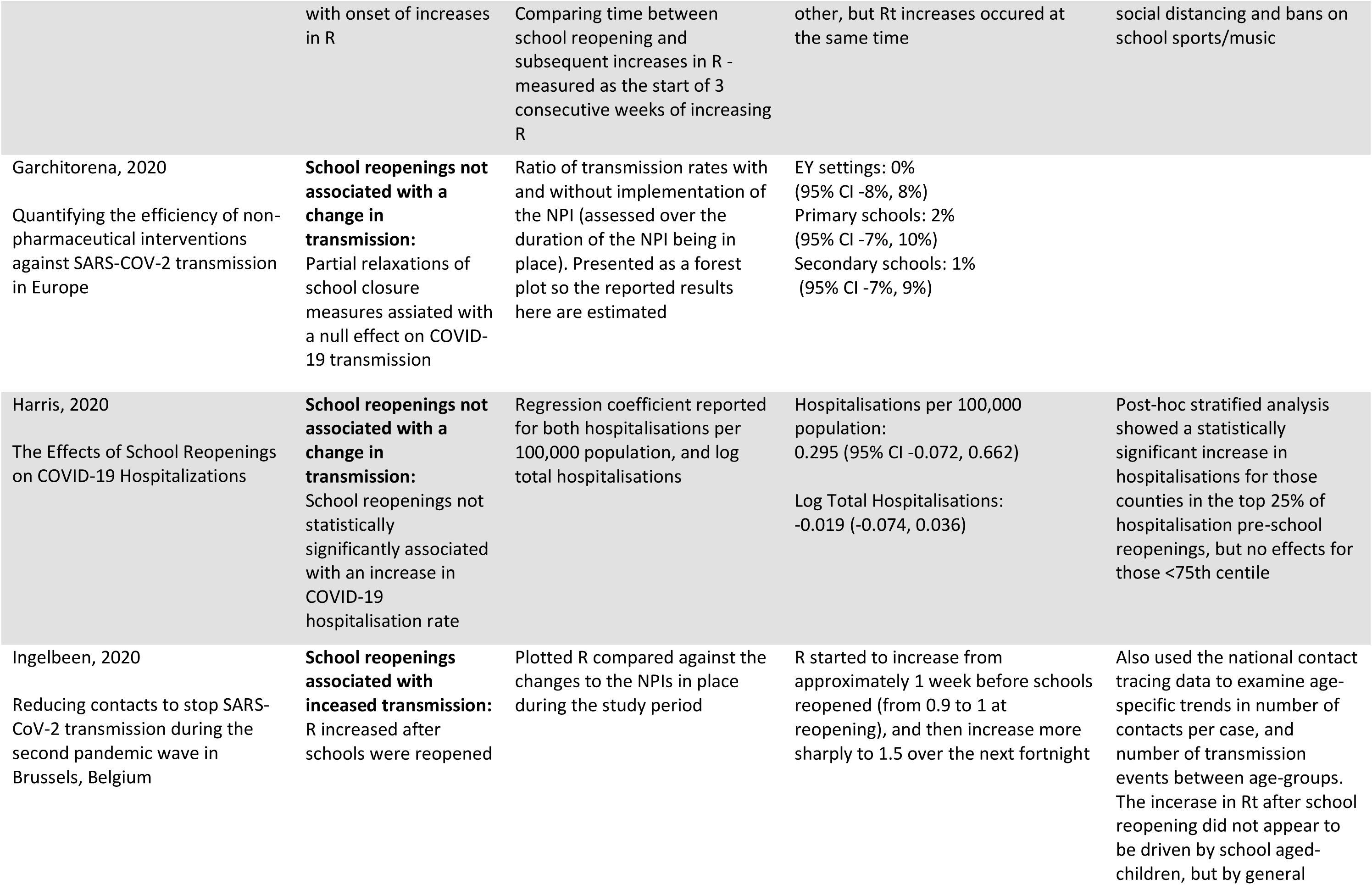
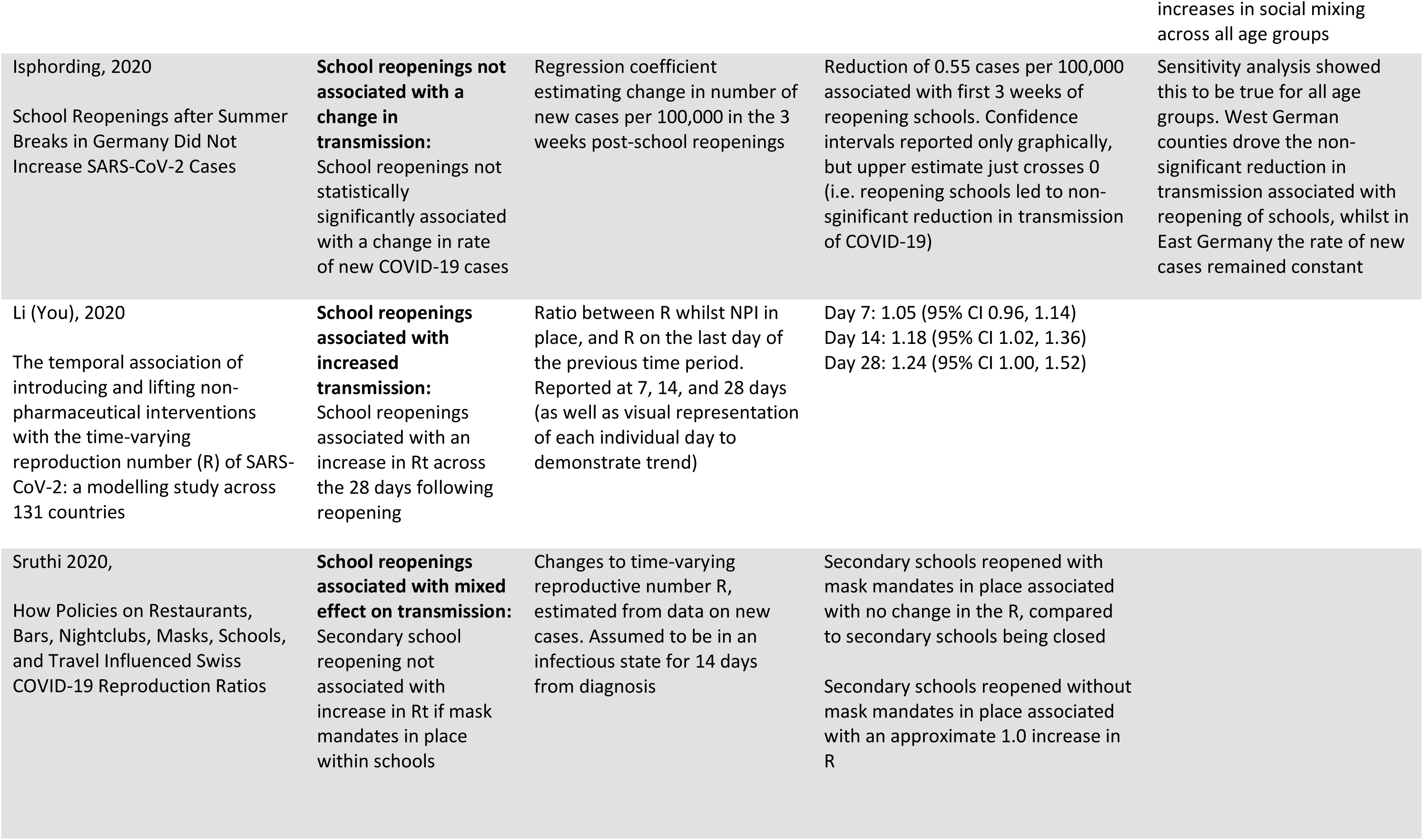
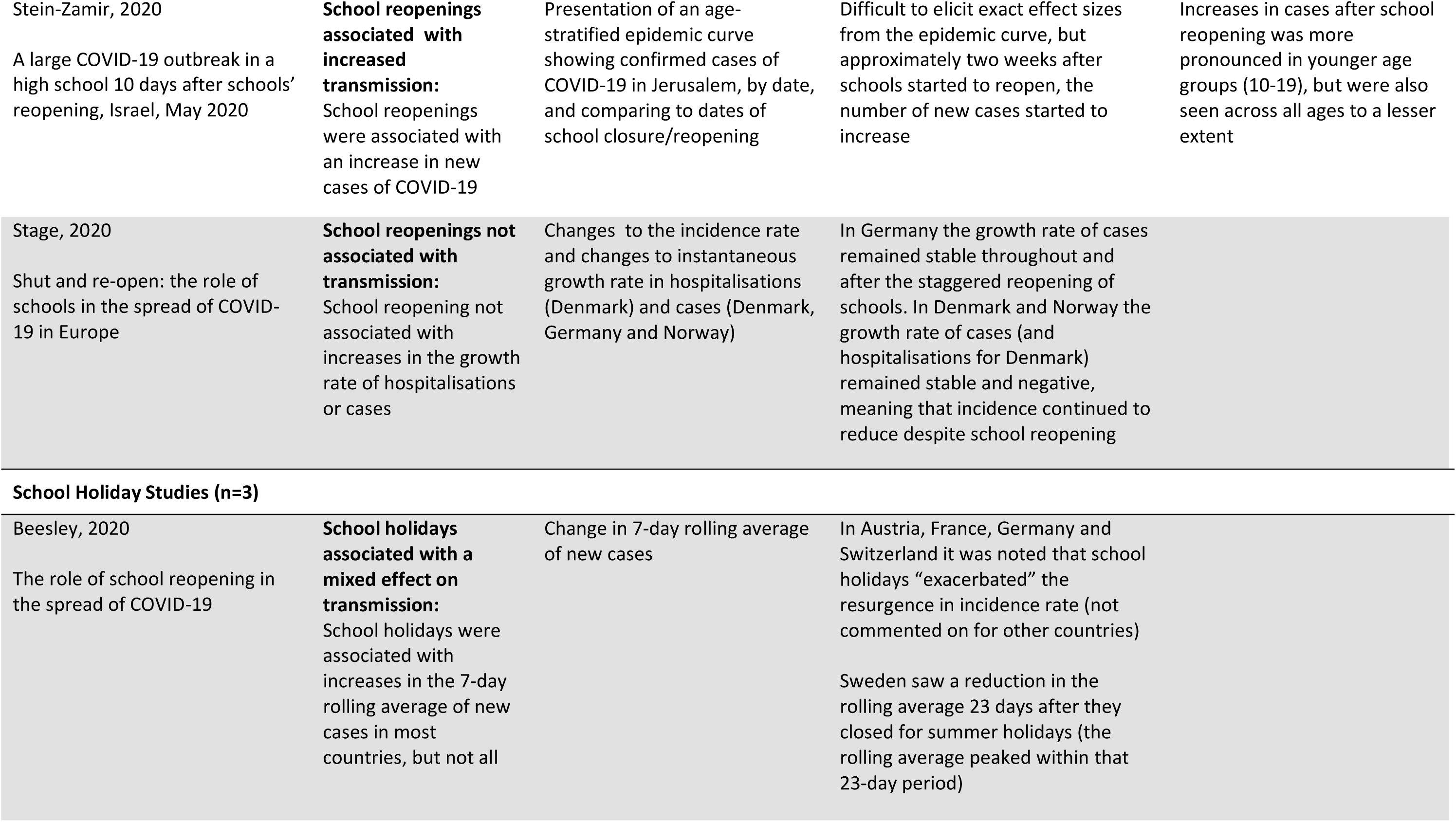
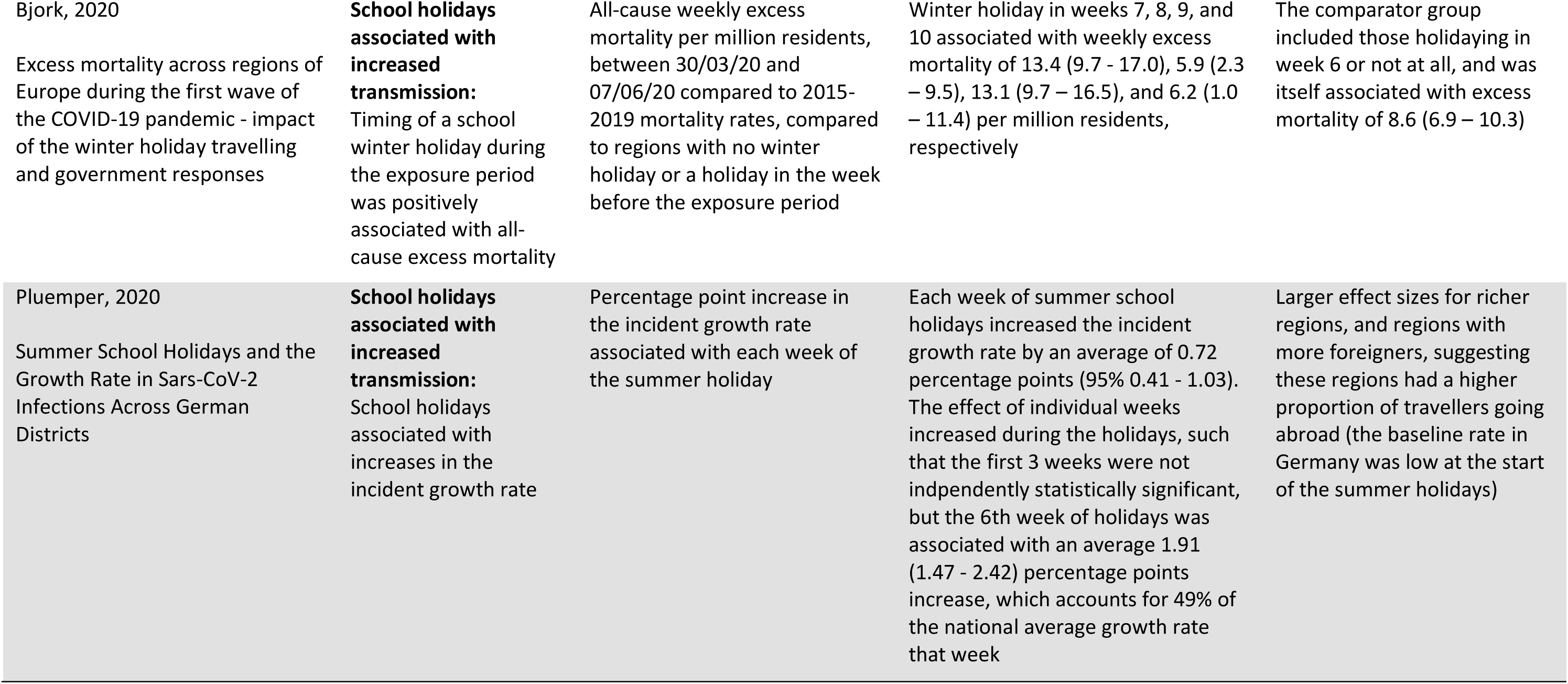
Findings from included studies, stratified by study design.

### Narrative Synthesis of Findings

#### School Closures

##### Pooled multiple-area before-after comparisons

We identified 22 studies(14,15,18–21,24,26,30,32–36,39,40,42,46–50) that analysed before-after data on multiple geographical units, and then pooled the results into one unified estimate of effect (generally by using regression analysis). These studies relied upon different timings of NPI implementation in different areas to establish their independent effects, and were therefore at risk of collinearity if compared areas implemented the same NPIs at similar times. These studies were also at risk of bias from sociocultural differences between compared areas.

Of these studies, 11(14,24,32,34–36,40,42,48–50) reported that school closures were associated with significantly reduced community transmission of SARS-CoV-2, seven(15,18,20,26,30,39,47) reported no association, and four(19,21,33,46) reported mixed findings. Those studies found to be at higher risk of bias, generally because they were judged not to have adjusted appropriately for NPIs, testing, or sociodemographic data, tended to report reductions in transmission; whereas those studies at lower risk of bias were as likely to report null effects as they were reductions (see figure 2).

Of the three studies(20)using this approach which were considered to be at the lowest risk of confounding, two reported no association and one reported that school closures reduced transmission. Courtemanche et al.(20) used a fixed effects model (to account for inter-area sociodemographic differences) in an event study design to estimate the effect of NPIs (including school closures) on SARS-CoV-2 incidence in US counties between March and April 2020. They adjusted for relevant NPIs, testing regime confounders, and underlying trends in each counties’ growth rates, and reported a null effect of school closures on growth rate, applying a lag of either 10 or 20 days. Hsiang et al.(26) used a reduced form of econometric regression to compare changes in incidence in French regions, Italian regions, and US states (in three separate analyses) before and after NPI implementation (including school closures) until early April 2020. Other key NPIs and testing regimes were adjusted for. The authors report a null effect of school closures on growth rate of SARS-CoV-2 incidence, with narrow confidence intervals for France and the USA, but a regression coefficient suggestive of a non-significant preventative effect in Italy (−0·11 (95% CI −0·25, 0·03)). Li et al.(54), used the’EpiForecast’ model of R(54) to estimate the effectiveness of different NPIs (including school closures) over time in 131 countries between January and June 2020. They identified time periods in which the NPIs in a given country were static, and calculated the ‘R ratio’ by dividing the average daily R of each period by the R from the last day of the previous period. They reported pooled estimates, regressed across all countries, for the first 28 days after introduction/relaxation of each NPI. Though the confidence intervals for each daily effect size included 1, the trend was clearly towards a reduction in transmission following school closure implementation.

##### Within-area before-after comparisons

We identified seven studies(23,29,37,38,43,44,53) that compared community transmission of SARS-CoV-2 before and after school closure for single geographical units, and did not pool the results with those of other areas. This approach controls for confounding from population sociodemographic factors, but remains vulnerable to confounding from other NPIs and temporal changes to testing regimes. As with the pooled before-after comparison studies, those studies at higher risk of bias from confounding were more likely to report reductions in transmission associated with school closures.

One study using this approach was found to be at moderate risk of bas. Matzinger et al.(37) identified the three US states which introduced school closures first, and with a sufficient lag before implementing other measures to assess their specific impact. They plotted incidence rates on a log_2_ scale and identified points of inflexion in the period after school closure. This assumes exponential growth in the absence of interventions, which may not have occurred given changes to testing regimes. The doubling time of new cases in Georgia slowed from 2·1 to 3·4 days one week after closing schools. Similar results were observed in Tennessee (2·0 to 4·2 days after one week) and Mississippi (1·4 to 3·4 days after two weeks). The authors also noted inflexion points for hospitalisations and mortality at later time points, although numerical changes were not reported. Tennessee showed a slowing in hospitalisations one week after cases, and mortality one week after hospitalisation. Mississippi showed a slowing in hospitalisations and mortality at the same time, one week after cases – the authors do not comment on this discrepancy. Georgia lacked early hospitalisation data to make such a comparison.

##### Pooled multiple-area comparisons of interventions in place at a fixed time point

Three studies(31,51,52) considered countries from around the world using a design in which NPIs were considered as binary variables on a specific date (i.e. in place or not in place), and the cumulative incidence or mortality to that point was compared to the number of new cases of COVID-19 over a subsequent follow-up period; countries were then compared using regression analysis to elicit independent effect sizes for individual policies including school closures. This approach reduces bias from different testing regimes over time and between countries. However, the use of a single cut-off date for whether school closure was in place means that that the effects of longer-standing and recent school closures were pooled, introducing misclassification bias. Two of these studies(31, 51) were at serious risk of bias and reported that school closures were associated with lower incidence; and one study(52) was at critical risk of bias and reported that closing schools was not associated with incidence but was associated with increased mortality. Each of these studies was at high risk of confounding from other NPIs, in addition to the risk of misclassification bias described above.

##### School Reopening Studies

Eleven studies(16,22–25,27,28,35,43–45) considered the effect of school reopening on subsequent SARS-CoV-2 community transmission(24). Of these, five were pooled multiple-area before-after comparison studies(24,25,28,35,43), and six were within-area multiple-area before-after comparison studies(16,22,23,27,44,45). These studies benefited from more staggered lifting of restrictions (compared to their implementation), and more stable testing regimes.

Of the four studies at a lower risk of bias(24,25,28,35), three(24,25,28) reported that schools were reopened without associated increases in transmission, whilst one(35) reported increased transmission. Garchitorena et al.(24) compared incidence data, with adjustment for underdetection, from 32 European countries, using multivariate linear regression models with adjustment for other NPIs and fixed effects to account for inter-country sociodemographic differences. They reported no association with incidence rates up to 16/09/20 of reopening early years settings (0% mean change in incidence rate (95% CI −8%, 8%)), primary schools (2% (−7%, 10%)), or secondary schools (−1% (−7%, 9%)). Harris et al.(25) estimated the effect of school reopenings on COVID-19 hospitalisation in the USA using an event study model, with analysis at the county-level. They adjusted for other NPIs, and used fixed effects to account for calendar week effects and inter-county differences. They applied a one week lag period, and compared data from ten weeks before, to six weeks after school reopenings. They initially report null effects when pooling the effects across all counties, however, post-hoc sensitivity analyses suggested that there were increases in hospitalisations for counties that were in the top 25% of baseline hospitalisation rate at school reopening (compared to null effects for the bottom 75%). Isphording et al.(28) compared changes to the COVID-19 incidence rate in German counties that were first to reopen schools after the summer holidays, with those yet to reopen (noting that the timing of such decisions was set years in advance, and not changed due to the pandemic). They considered data from two weeks before to three weeks after school reopenings, and adjusted for mobility data, and used fixed effects to account for inter-county sociodemographic differences. They reported no association between school reopenings and incidence. One study, by You Li et al.(35), is described above as it reports on the effect of both school closures and school reopenings around the World. As for school closures, their effect sizes for each individual day in the 28-day period post-school reopenings were not always statistically significant, but the data trend is clearly that of an increase in transmission associated with school reopenings.

The seven studies(16,22,23,27,43–45) at serious and critical risk of confounding are more difficult to interpret, again predominantly due to the high risk of confounding. Three(16,23,44) reported no association between school reopening and transmission, two(22, 43) reported mixed findings, and two(27, 45) reported increased transmission following reopening of schools.

##### School Holiday Studies

Three studies(16,17,41) reported changes in SARS-CoV-2 community transmission associated with school holidays. These holidays occurred according to pre-determined timetables and are therefore unlikely to be influenced by background trends in infections. Two studies examined associations between timing of summer holidays on incidence rates in Germany(41) and in multiple European countries(16), respectively. The other study(17) reported on the timing of the February/March 2020 half-term break timing in countries that neighbour the Alps. Of these, one reported mixed findings on the effect of summer holidays(16), and two reported that school holidays were associated with increased transmission(17, 41). The authors of these studies considered the primary exposure to be increased social contact from international travel, rather than decreases from the temporary closure or schools.

##### Different School Setting Types

One school closure study(48), three school reopening studies (16,22,44), and one study looking at closures and reopenings(24) considered evidence of independent effects for different types of school closures.

Two studies reported independent effect sizes for different settings, but found considerable overlap between the effect sizes, and noted high temporal correlation between the policy timings meaning that collinearity limits the interpretability of the findings. Garchitorena et al.(24) (moderate risk of bias) reported the effect of both school closures and school reopenings on changes to R in 32 European countries, with almost completely overlapping estimates of transmission reductions associated with closures in early years settings, primary schools and secondary schools; and equally null effects for each setting associated with reopenings. Yang et al.(48) (moderate risk of bias) reported that school closures in US counties (presumed primary and secondary combined) were associated with 37% (95% CI 33-40%) reductions in R, compared to 31% reductions for early years settings (95% CI 26-35%).

Two studies reported staggered reopenings of different school settings, generally with younger children students returning first, and a week or two between each reopenings. Both studies found null effects on transmission overall, and therefore did not report any differential effect by setting type. Stage et al.(44) (serious risk of bias) noted staggered reopenings in Norway, Denmark and Germany. Ehrhardt et al.(22) (critical risk of bias) noted staggered reopenings of schools in Baden-Wuttemberg (a region of Germany)

Beesley(16) (critical risk of bias) noted that increases in the 7-day rolling average of new cases were greater in the 40 days after secondary school reopening than they were in the 24 days following primary schools reopening. However, this study is at high risk of confounding from other NPIs, and it is not clear why the chosen (and different) lag periods were applied.

## Discussion

We identified 40 studies that provided a quantitative estimate of the impact of school closures or reopening on community transmission of SARS-CoV-2. The studies included a range of countries and were heterogenous in design. Amongst higher quality, less confounded studies of school closures, 6 out of 14 reported that school closures had no effect on transmission, 6 reported that school closures were associated with reductions in transmission, and 2 reported mixed findings (figure 2); with findings ranging from no association to a 60% relative reduction in incidence and mortality rate(14). Most studies of school reopening reported that school reopening, with extensive infection prevention and control measures in place and when the community infection levels were low, did not increase community transmission of SARS-CoV-2

The strength of this study is that it draws on empirical data from actual school closures and reopenings during the COVID-19 pandemic and includes data from 150 countries. By necessity, we include observational rather than randomised controlled studies, as understandably no jurisdictions have undertaken such trials. We were unable to meta-analyse due to study heterogeneity. We were unable to meaningfully examine differences between primary and secondary schools as very few studies distinguished between them, despite the different transmission patterns for younger and older children. Data are also lacking from low-income countries, where sociocultural factors may produce different effects of school closures on transmission to high income settings, leaving a substantial gap in the evidence base. Data in these studies comes exclusively from 2020, and many studies report only up to the summer months, it is therefore unclear whether our findings are robust to the effects of new SARS-CoV-2 variants and vaccines.

A major challenge with estimating the ‘independent’ effect of school closures, acknowledged by many of the studies, is disentangling their effect from other NPIs occurring at the same time. While most studies tried to account for this, it is unclear how effective these methods were. Even where adjustment occurred there is a risk of residual confounding, which likely overestimated preventative associations; and collinearity (highly-correlated independent variables meaning that is impossible to estimate specific effects for each) which could bias results towards or away from the null. One exception was a paper by Matzinger et al.(37) which focused on three US states that implemented school closures first and without co-interventions, and reported a two-fold increase in the time for cases to double one week after school closures. However it is possible that the benefits observed here may be attributable, at least in part, to a ‘signalling effect’ with other changes to social mobility (e.g. working from home) being prompted by school closures. Another approach, though ineligible for inclusion in our study, is to examine transmission data for breakpoints, and then work backwards to see what NPIs were in place at the time. Two studies that did this found that transmission started to drop following other NPIs, before school closures were implemented, and found no change in the gradient of decline after school closures in Switzerland(55) and Germany(56). This may suggest school closures have different effects when implemented first, or on top of other restrictions, perhaps due to a broader signalling effect that the first implemented NPI has on societal mobility patterns. The true independent effect of school closures from the first wave around the world may simply be unknowable.

In contrast, lifting of NPIs in the summer of 2020 (including school reopenings) generally occurred in a more staggered way, and on a background of stable testing regimes and outcome ascertainment. Good-quality observational studies considering data from across 32 European countries(24), Germany alone(28), and the USA(25) all demonstrated that school reopenings can be successfully implemented without increasing community transmission of SARS-CoV-2, where baseline incidence is low and robust infection prevention and control measures are in place. However, the USA-based study did comment that those counties with the highest 25% of baseline hospitalisations at the time of reopenings (above 40 admissions per 100,000 population per week) did see an increase in transmission following school reopenings, although the bottom 75% of counties did not see any effect. This may explain why the other school reopening study at lower risk of bias(35) reported a clear, though non-significant, trend towards school reopenings being associated with increases in transmission rates across 131 countries worldwide, with the authors noting “we were unable to account for different precautions regarding school reopening that were adopted by some countries” before citing Israel as an example where an uptick in transmission occurred following reopening, and where “students were in crowded classrooms and were not instructed to wear face masks.”

The variability in findings from our included studies are likely to reflect issues with study design. However, this may also suggest that there is no single effect of school closures and reopenings on community transmission and that contextual factors modify the impact of closures in different countries and over time. If the purpose of school closures is reduction in social contacts among children, the level of social mixing between children that occurs outside school once schools are closed is likely to be a key determinant of their effect at reducing community transmission. This will be influenced by other NPIs, and other key contextual factors including background prevalence of infection, use of preventive measures in schools prior to closures, age of children affected, as well as sociodemographic and cultural factors.

Different countries have adopted different approaches to controlling COVID-19. Early in the pandemic school closures were common, and in some places were one of the first major social distancing measures introduced. The effectiveness of the overall bundle of lockdown measures implemented is proven, but the incremental benefit of school closures remains unclear. In contrast, only one of the four studies of school reopenings assessed at a lower risk of bias reported an increase in community transmission. Collectively the evidence around school re-openings, while more limited in size, tends to suggest that school reopenings, when implemented during periods of low incidence and accompanied by robust preventive measures, are unlikely to have a measurable impact on community transmission. Further research is needed to validate these findings and their generalisability, including with respect to new variants. These findings are highly important given the harmful effects of school closures(3, 4). Policymakers and governments need to take a measured approach before implementing school closures in response to rising infection rates, and look to reopen schools, with appropriate mitigation measures in place, where other lockdown measures have successfully brought community transmission of SARS-CoV-2 under control.

## Supporting information

PRISMA Checklist

## Data Availability

All included articles are publicly available.

## Author Contributions

SW, CW, CBo, RV and OM designed the review protocol. SW, AC, VB, SR and JB screened articles for inclusion, assessed risk of bias, and performed data extraction. SW and OM drafted the manuscript. All authors commented on the final manuscript.

## Appendix A Search Strategy

**Search dates: 12/10/20 and 07/01/21**

### PubMed

Search Title/Abstract:

(coronavirus[mh] OR Coronavirus Infections[mh] OR coronavirus*[tw] OR “COVID-19”[tw] or “2019-nCoV”[tw] or “SARS-CoV-2”[tw]) AND (Schools[mh:noexp] OR schools, nursery[mh] OR “Child Day Care Centers”[mh] OR “Nurseries, Infant”[mh] OR school*[tiab] OR preschool*[tiab] OR “pre-school*”[tiab] OR nurser*[tiab] OR kindergarten*[tiab] OR “day care”[tiab] OR daycare[tiab] OR “education setting*”[tiab] OR “educational setting*”[tiab] OR NPI*[tiab] OR “non-pharmaceutical intervention*”[tiab])

### Web of Science

TS=(coronavirus* OR “COVID-19” OR “2019-nCoV” OR “SARS-CoV-2”)

AND

TS=(school* OR nurser* OR preschool* OR “pre-school*” OR kindergarten* OR “day care” OR

daycare OR “education setting*” OR “educational setting*” OR NPI* OR “non-pharmaceutical intervention*”)

### Scopus

TITLE-ABS-KEY ((coronavirus* OR “COVID-19” OR “2019-nCoV” OR “SARS-CoV-2”) AND (school* OR nurser* OR preschool* OR “pre-school*” OR kindergarten* OR “day care” OR “daycare” OR “education setting*” OR “educational setting*” OR NPI* OR “non-pharmaceutical intervention*”)) AND (LIMIT-TO (PUBYEAR, 2020))

### CINAHL (via HDAS)

((coronavirus* OR “COVID-19” OR “2019-nCoV” OR “SARS-CoV-2”) AND (school* OR nurser* OR preschool* OR “pre-school*” OR kindergarten* OR “day care” OR “daycare” OR “education setting*” OR “educational setting*” OR NPI* OR “non-pharmaceutical intervention*”)).ti,ab [DT 2020-2020]

### WHO Global COVID-19 Research Database

(tw:(school*)) OR (tw:(nurser*)) OR (tw:(“pre-school*”)) OR (tw:(preschool*) OR (tw:(kindergarten*)) OR tw:(“day care”) OR tw:(“daycare”) OR tw:(“education setting*”) OR tw:(“educational setting*”) OR tw:(NPI*) OR tw:(“non-pharmaceutical intervention*”))

Including: WHO COVID Database, MedRxiv. Title, abstract, subject. 2020.

### ERIC

Coronavirus OR “COVID-19” or “2019-nCoV” or “SARS-CoV-2”

### British Education Index

Coronavirus OR “COVID-19” or “2019-nCoV” or “SARS-CoV-2”

### Australian Education Index

Coronavirus OR “COVID-19” or “2019-nCoV” or “SARS-CoV-2”

### Grey Literature Search, Google

First 100 hits on google search, limiting to PDF files, up to ‘last year’.

Search: “COVID-19” OR “coronavirus” OR “school” OR “education”

