## Supplementary material for "Do school closures and school reopenings affect community transmission of COVID-19? A systematic review of observational studies": PRISMA Checklist

### PRISMA 2009 Checklist

| Section/topic | # | Checklist item | Reported on page # |
|---|---|---|---|
| **TITLE** | | | |
| Title | 1 | Identify the report as a systematic review, meta-analysis, or both. | Title |
| **ABSTRACT** | | | |
| Structured summary | 2 | Provide a structured summary including, as applicable: background; objectives; data sources; study eligibility criteria, participants, and interventions; study appraisal and synthesis methods; results; limitations; conclusions and implications of key findings; systematic review registration number. | Page 1 |
| **INTRODUCTION** | | | |
| Rationale | 3 | Describe the rationale for the review in the context of what is already known. | Page 3: School closures have been a common strategy to control the spread of SARS-CoV-2…school closures have significant negative consequences…the specific contribution of school closures [to limiting SARS-CoV-2 spread] remains unclear |
| Objectives | 4 | Provide an explicit statement of questions being addressed with reference to participants, interventions, comparisons, outcomes, and study design (PICOS). | Page 3: Here, we synthesise the observational evidence of the impact of closing or reopening schools on community transmission of SARS-CoV-2. |
| Protocol and registration | 5 | Indicate if a review protocol exists, if and where it can be accessed (e.g., Web address), and, if available, provide registration information including registration number. | Page 5: Prospero (ID:CRD42020213699). |
| Eligibility criteria | 6 | Specify study characteristics (e.g., PICOS, length of follow-up) and report characteristics (e.g., years considered, language, publication status) used as criteria for eligibility, giving rationale. | Page 5: We included any empirical study which reported a quantitative estimate of the effect of school closure or reopening on community transmission of SARS-CoV-2. We considered 'school' to include early years settings (e.g. nurseries or kindergartens), primary schools, and secondary school, but excluded further or higher education (e.g. universities). Community transmission was defined as any measure of community infections rate, hospital admissions, or mortality attributed to COVID-19. We included studies published in 2020 or 2021 only. We included pre-prints, peer-reviewed and grey literature. We did not apply any restriction on language, but all searches were undertaken in English. We excluded prospective modelling studies and studies in which the assessed outcome was exclusively transmission within the school environment rather than the wider community. |

| | | | |
|---|---|---|---|
| Information sources | 7 | Describe all information sources (e.g., databases with dates of coverage, contact with study authors to identify additional studies) in the search and date last searched. | Page 5: We searched PubMed, Web of Science, Scopus, CINAHL, the WHO Global COVID-19 Research Database (including medRxiv and SSRN), ERIC, the British Education Index, and the Australian Education Index, searching title and abstracts for terms related to SARS-CoV-2 AND terms related to schools or NPIs. To search the grey literature, we searched Google. We also included papers identified through professional networks. Full details of the search strategy are included in Appendix A. Searches were undertaken first on 12 October 2020 and updated on 07 January 2021. |
| Search | 8 | Present full electronic search strategy for at least one database, including any limits used, such that it could be repeated. | Appendix A includes full search strategy |
| Study selection | 9 | State the process for selecting studies (i.e., screening, eligibility, included in systematic review, and, if applicable, included in the meta-analysis). | Page 6: Article titles and abstracts were imported into the Rayyan QCRI webtool(11). Two reviewers independently screened titles and abstracts, retrieved full texts of potentially relevant articles, and assessed eligibility for inclusion. |
| Data collection process | 10 | Describe method of data extraction from reports (e.g., piloted forms, independently, in duplicate) and any processes for obtaining and confirming data from investigators. | Page 6: Two reviewers independently extracted data and assessed risk of bias. Data extraction was performed using a pre-agreed extraction template which collected information on publication type (peer-reviewed or pre-print), country, study design, exposure type (school closure or re-opening), setting type (primary or secondary), study period, unit of observation, confounders adjusted for, other NPIs in place, analysis method, outcome measure, and findings. |
| Data items | 11 | List and define all variables for which data were sought (e.g., PICOS, funding sources) and any assumptions and simplifications made. | Page 6: Two reviewers independently extracted data and assessed risk of bias. Data extraction was performed using a pre-agreed extraction template which collected information on publication type (peer-reviewed or pre-print), country, study design, exposure type (school closure or re-opening), setting type (primary or secondary), study period, unit of observation, confounders adjusted for, other NPIs in place, analysis method, outcome measure, and findings. |
| Risk of bias in individual studies | 12 | Describe methods used for assessing risk of bias of individual studies (including specification of whether this was done at the study or outcome level), and how this information is to be used in any data synthesis. | Page 6: We used the Cochrane Risk of Bias In Non-randomised Studies of Interventions (ROBINS-I) tool(12) to evaluate bias. |
| Summary measures | 13 | State the principal summary measures (e.g., risk ratio, difference in means). | Page 6: Community transmission was defined as any measure of community infection rate, hospital admission rate, or mortality attributed to COVID-19. |
| Synthesis of results | 14 | Describe the methods of handling data and combining results of studies, if done, including measures of consistency (e.g., $I^2$) for each meta-analysis. | Page 6: Given the heterogeneous nature of the studies, prohibiting meta-analysis, a narrative synthesis was conducted. |

### PRISMA 2009 Checklist

| Section/topic | # | Checklist item | Reported on page # |
|---|---|---|---|
| Risk of bias across studies | 15 | Specify any assessment of risk of bias that may affect the cumulative evidence (e.g., publication bias, selective reporting within studies). | n/a |
| Additional analyses | 16 | Describe methods of additional analyses (e.g., sensitivity or subgroup analyses, meta-regression), if done, indicating which were pre-specified. | n/a |
| **RESULTS** | | | |
| Study selection | 17 | Give numbers of studies screened, assessed for eligibility, and included in the review, with reasons for exclusions at each stage, ideally with a flow diagram. | Page 7: We identified 7,474 studies (Figure 1). After removing 2,339 duplicates, 5,135 unique records were screened for inclusion. We excluded 4,842 records at the title or abstract stage, leaving 293 records for full text review. Of these, 40(14–53) met the inclusion criteria. |
| Study characteristics | 18 | For each study, present characteristics for which data were extracted (e.g., study size, PICOS, follow-up period) and provide the citations. | Table 1 |
| Risk of bias within studies | 19 | Present data on risk of bias of each study and, if available, any outcome level assessment (see item 12). | Table 2 |
| Results of individual studies | 20 | For all outcomes considered (benefits or harms), present, for each study: (a) simple summary data for each intervention group (b) effect estimates and confidence intervals, ideally with a forest plot. | Table 3 |
| Synthesis of results | 21 | Present results of each meta-analysis done, including confidence intervals and measures of consistency. | n/a |
| Risk of bias across studies | 22 | Present results of any assessment of risk of bias across studies (see Item 15). | Figure 2 |
| Additional analysis | 23 | Give results of additional analyses, if done (e.g., sensitivity or subgroup analyses, meta-regression [see Item 16]). | n/a |
| **DISCUSSION** | | | |
| Summary of evidence | 24 | Summarize the main findings including the strength of evidence for each main outcome; consider their relevance to key groups (e.g., healthcare providers, users, and policy makers). | Page 16: Amongst higher quality, less confounded studies of school closures, 6 out of 14 reported that school closures had no effect on transmission, 6 reported that school closures were associated with reductions in transmission, and 2 reported mixed findings (figure 2); with findings ranging from no association to a 60% relative reduction in incidence and mortality rate(14). Most studies of school reopening reported that school reopening, with extensive infection prevention and control measures in place and when the community infection |
